# Heterogeneous aging across multiple organ systems and prediction of chronic disease and mortality

**DOI:** 10.1101/2022.09.03.22279337

**Authors:** Ye Ella Tian, Vanessa Cropley, Andrea B. Maier, Nicola T. Lautenschlager, Michael Breakspear, Andrew Zalesky

## Abstract

Biological aging of human organ systems reflects the interplay of age, chronic disease, lifestyle and genetic risk. Using longitudinal brain imaging and physiological phenotypes from the UK Biobank, we establish normative models of biological age for 3 brain and 7 body systems. We find that an organ’s biological age selectively influences the aging of other organ systems, revealing a multiorgan aging network. We report organ age profiles for 16 chronic diseases, where advanced biological aging extends from the organ of primary disease to multiple systems. Advanced body age associates with several lifestyle and environmental factors, leucocyte telomere lengths and mortality risk, and predicts survival time (AUC=0.77) and premature death (AUC=0.86). Our work reveals the multisystem nature of human aging in health and chronic disease. It may enable early identification of individuals at increased risk of aging-related morbidity and inform new strategies to potentially limit organ-specific aging in such individuals.

## Introduction

Age is the greatest common risk factor for chronic diseases^1, 2^. However, trajectories of age-related decline vary markedly between individuals and differ across human organ systems^3, 4^. Biological age is thus recognized as a more informative marker of disease risk and mortality than chronological age^5, 6^. As a result, cellular, molecular and physiological aging biomarkers^7, 8^ have been developed and studied across multiple species ^9–14^.

To fulfill the clinical potential of this work, biological aging clocks that are specific to particular organ systems, tissue types and aging-related diseases are now required to be established in large and diverse longitudinal populations^15^. A multiorgan characterization of biological aging across major chronic dieases can facilitate novel organ-specific therapeutic opportunities, yield disease-specific risk calculators and elucidate factors that drive the divergence of an organ’s biological age from chronological age. Elucidating such factors will inform strategies to potentially slow age-related decline, reduce the risk of chronic diseases and promote healthy longevity^16–19^.

Biological aging of the human brain has been the focus of considerable research^20–24^. Predictive models of brain age derived from neuroimaging can infer apparent age based on brain structure and function. The difference between chronological age and predicted brain age, known as the *brain age gap*, provides a measure of biological age and can reveal insight into whether an individual’s brain appears older or younger relative to same-aged peers^20^. While age gaps may first emerge in early life and accumulate across the lifespan, longitudinal increases in age gaps later in life relate more specifically to aging-related decline. In principle, biological age can be estimated *in vivo* for organs and body systems other than the brain. Organ-specific age gaps will enable concurrent investigation of biological aging across multiple body and brain systems. To this end, we develop new assays to measure the biological age (i.e., age gap index) for 7 body and 3 brain systems using imaging, physiological and blood-derived phenotypes acquired cross-sectionally (body: n=143,423; brain: n=36,901) and longitudinally (body: n=1,220; brain: n=1,294) in the UK Biobank cohort. We aim to: (i) map the influence of an organ’s biological age on the aging of other organ systems; (ii) elucidate body and brain age profiles characteristic of 16 aging-related chronic diseases; (iii) establish whether organ-specific biological age associates with lifestyle factors and leucocyte telomere length; and (iv) predict the risk of mortality using body and brain age profiles. Our work reveals the heterogeneity of biological aging across individuals and organs, and its relation to lifestyle factors, risk of specific chronic diseases and mortality in midlife and older adults. Quantifying the impact of major chronic diseases on organ aging holds substantial promise for precision geriatric medicine and related clinical translation.

## Results

Multimodal brain imaging, physiological and blood phenotypes were grouped based on their relevance to the structure and function of specific organ systems; namely, 7 body (cardiovascular, pulmonary, musculoskeletal, immune, renal, hepatic and metabolic; table S1) and 3 brain (gray matter, white matter and brain connectivity; table S2) systems. Cognitive performance formed an additional group (table S3). Phenotypes for body systems were available for 143,423 individuals (age range 39-73 years, mean 56.7±8.2, 79,980 males), and brain phenotypes were available for 36,901 individuals (age range 45-82 years, mean 64.2±7.5, 17,203 males). Cognitive phenotypes were available for 32,317 individuals (age range 45-82 years, mean 65.1 ± 7.6, 15,712 males).

Healthy adults with no major medical conditions were selected to train machine learning models to predict individual chronological age (see Methods). Separate predictive models of chronological age were established for each body/brain system and sex. Subtracting actual chronological age from predicted chronological age, referred to as the *age gap*, captures whether an individual’s organ system appears older (gap>0) or younger (gap<0) than population norms for the individual’s chronological age and sex. Age gaps thus provide normative, organ-specific clocks of biological age (Fig. 1).

**Fig. 1.**
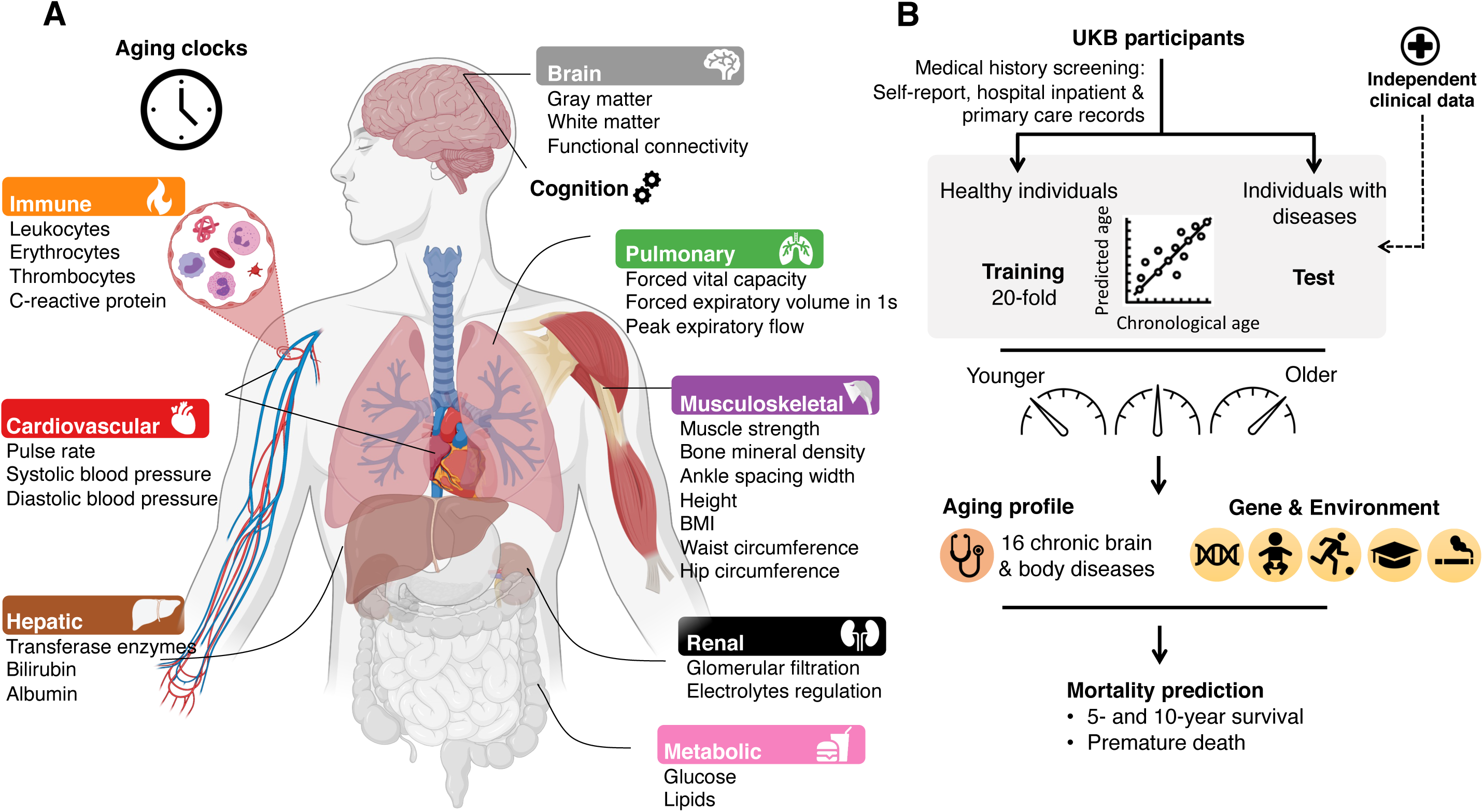
Overview of study design. (**A**) Organ systems for which normative models of biological age were established using organ-specific phenotypes. Key phenotypes are listed below each system. Image was created with BioRender.com. (**B**) Predictive models of chronological age were established using phenotypes from healthy adults and 20-fold cross-validation. Separate models were developed for each body/brain system and sex. Using models trained on healthy adults, personalized body and brain age gaps were determined for individuals with lifetime diagnoses of chronic diseases to investigate the relation of biological age with disease and mortality. Two independent datasets were used to validate brain age gap estimates in individuals with dementia. Associations between genetic/environmental factors and biological age were also investigated.

### Normative aging models

Chronological age could be predicted with modest to high accuracy for body (female: r=0.79, mean absolute error (MAE)=3.71 years; male: r=0.72, MAE=4.46 years) and brain (female: r=0.79, MAE=3.52 years; male: r=0.80, MAE=3.68 years) systems and cognition (female: r=0.53, MAE=4.87 years; male: r=0.54, MAE=5.21 years, Fig. 2A). Prediction accuracies varied between organ systems and sexes (Fig. 2B & fig. S1 & table S4). Comparable accuracies for brain systems were achieved in additional datasets (female: r=0.82, MAE=3.52; male: r=0.85, MAE=3.41, fig. S2A). Applying the trained models to predict the chronological age of all participants resulted in personalized organ-specific age gaps.

**Fig. 2.**
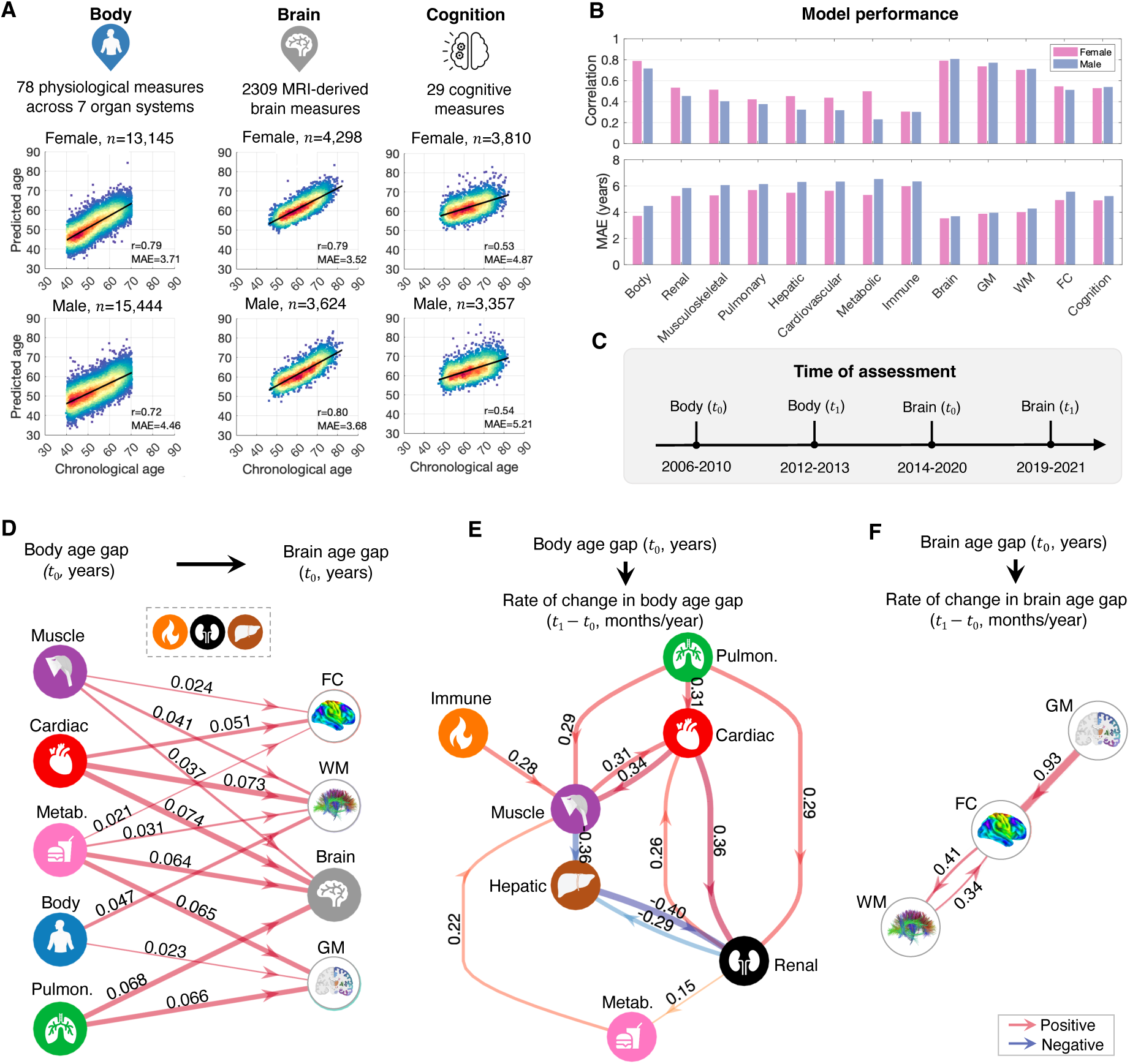
Age prediction accuracy and multiorgan aging networks. (**A**) Scatter plots show associations between chronological and predicted age for prediction models based on body (left), brain (mid) and cognitive (right) phenotypes. Lines of best fit indicated with solid black lines. n: training sample size; r: Pearson correlation coefficient; MAE: mean absolute error. (**B**) Bar plots show Pearson correlation coefficients (upper) and MAE (lower) quantifying age prediction accuracy (average of 10 repartitions of 20-fold cross-validation). (**C**) Assessment timeline for measures of body and brain function. (**D**) Influence of body age gaps (left) on brain age gaps (right), adjusting for sex, chronological age and the time interval between assessments. Links are shown for significant influences inferred from structural equation modeling (false discovery rate (FDR) corrected for 8 body ages × 4 brain ages = 32 tests). (**E**) Influence of baseline organ age on longitudinal rate of change in organ age, adjusting for overall body age gap, sex and chronological age at baseline. An arrow from organ *X* to organ *Y* indicates that the age gap of *X* at baseline significantly influences the rate of aging of *Y* (p<0.05, FDR corrected for 7 × 6 = 42 tests). (**F**) Same as panel (E) but for brain systems (p<0.05, FDR corrected for 3 × 2 = 6 tests). Edge thickness and color reflect regression coefficients (𝛽) estimated for edges comprising the structural equation model. Metab., metabolism; Pulmon., pulmonary; Muscle, musculoskeletal; Cardiac, cardiovascular; GM, gray matter; WM, white matter; FC, functional connectivity.

Follow-up phenotype and imaging measurements were available for body (n=1,220, 837 males; 2.1-5.6 years follow-up) and brain (n=1,294, 632 males; 2.0-2.7 years follow-up) systems. Chronological age was thus predicted at baseline (*t*_0_) and follow-up (*t*_1_), yielding two age gaps for each organ per individual (Fig. 2C). This enabled estimation of *longitudinal* rates of change in body and brain age.

### Multiorgan aging networks

Given that organ systems dynamically interact via nervous, circulatory and lymphatic networks^25^, we hypothesized that an organ’s age would selectively influence the rate of aging of several connected organ systems. Using structural equation modeling (SEM) on organ age gaps, we found that advanced biological age of several body systems explains advanced brain age (Fig. 2D). While these aging pathways are not necessarily causal in the strict sense, they reveal directional relationships elucidated through an established process of casual structure discovery (see Methods). For example, cardiovascular age demonstrates the strongest influence on brain age, where a one-year increase in cardiovascular age explains a 0.074 year (i.e., 27 day) increase in overall brain age, and 19 and 27 day increases in functional connectivity and white matter ages, respectively.

We next tested whether an organ’s baseline biological age influences the *rate of change* in the biological age of other organs. A positive influence would provide evidence consistent with *faster* aging^26^. Due to minimal overlap between individuals (n=17) with both longitudinal body and brain phenotypes, analyses were conducted separately for body and brain systems. The influence of one organ’s age gap on the rate of change in the age gap of each other organ was modeled using SEM (see Methods), yielding multiorgan aging networks (body: Fig. 2E, brain: Fig. 2F). The networks reveal several putative aging pathways. For example, advanced age of the pulmonary system leads to faster cardiovascular aging, which in turn results in faster aging of the musculoskeletal and renal systems (Fig. 2E). The cardiovascular-renal-metabolic-musculoskeletal systems form positive feedback loops, where faster aging is reinforced between organ systems. The musculoskeletal system is an in-degree hub, suggesting that faster musculoskeletal aging is a common sequela of aging across multiple organ systems (Fig. 2E).

For the brain, advanced gray matter age leads to faster aging of functional brain connectivity (i.e., each one-year increase in gray matter age at baseline leads to 28 days/year increase in the rate of aging of functional connectivity), but not the converse. A positive feedback loop is evident between functional connectivity and white matter (Fig. 2F). Including baseline and follow-up cognitive age gaps in the SEM did not reveal significant influences of baseline tissue-specific brain age on the rate of cognitive aging. Patterns of interorgan synchrony in biological age are shown in fig. S3 and SEM estimates are provided in table S5.

### Genetic, environmental and lifestyle associations with biological organ age

We next investigated genetic and environmental factors associated with organ age. Partial correlations were used to test for associations between organ-specific age gaps and 158 environmental/lifestyle measures, leucocyte telomere length and polygenic scores indexing leucocyte telomere length (see Methods). Several environmental and lifestyle factors explain significant variation in the biological age of multiple organs (p<2.6×10^-5^, Bonferroni corrected for 158 factors × 12 organ systems = 1896 tests, table S6). For most body systems (Fig. 3), individuals appearing older than same-aged peers were more likely to have smoked tobacco, consumed more alcohol and experienced long-standing illness, had menopause early in life and lived in areas of greater socioeconomic inequality. In contrast, those who exercised (faster than usual walking pace), had a larger birth weight, completed tertiary education and were older at first live birth were more likely to appear younger. Some lifestyle factors exclusively associate with organ-specific age gaps. For example, advanced pulmonary system age associates with exposure to air pollution, but not natural/green environments. Advanced brain age most strongly associates with smoking, alcohol consumption, long-standing illness and hearing loss. Interestingly, advanced cognitive age not only significantly associates with advanced brain age, but also with advanced age of several body systems, including pulmonary and musculoskeletal systems. Associations with tissue-specific brain age are shown in fig. S4. Shorter leucocyte telomere lengths weakly associate with older body (r=-0.033, p=2.5×10^-34^), pulmonary (r=-0.023, p=1.1×10^-17^), immune (r=-0.037, p=2.4×10^-42^) and renal (r=-0.02, p=7.3×10^-14^) age gaps. Similarly, polygenic scores indexing leucocyte telomere length weakly associate with cardiovascular (r=0.014, p=2.7×10^-7^), pulmonary (r=0.016, p=1.5×10^-9^), immune (r=-0.015, p<2.0×10^-8^) and renal (r=-0.012, p=1.7×10^-5^) age gaps.

**Fig. 3.**
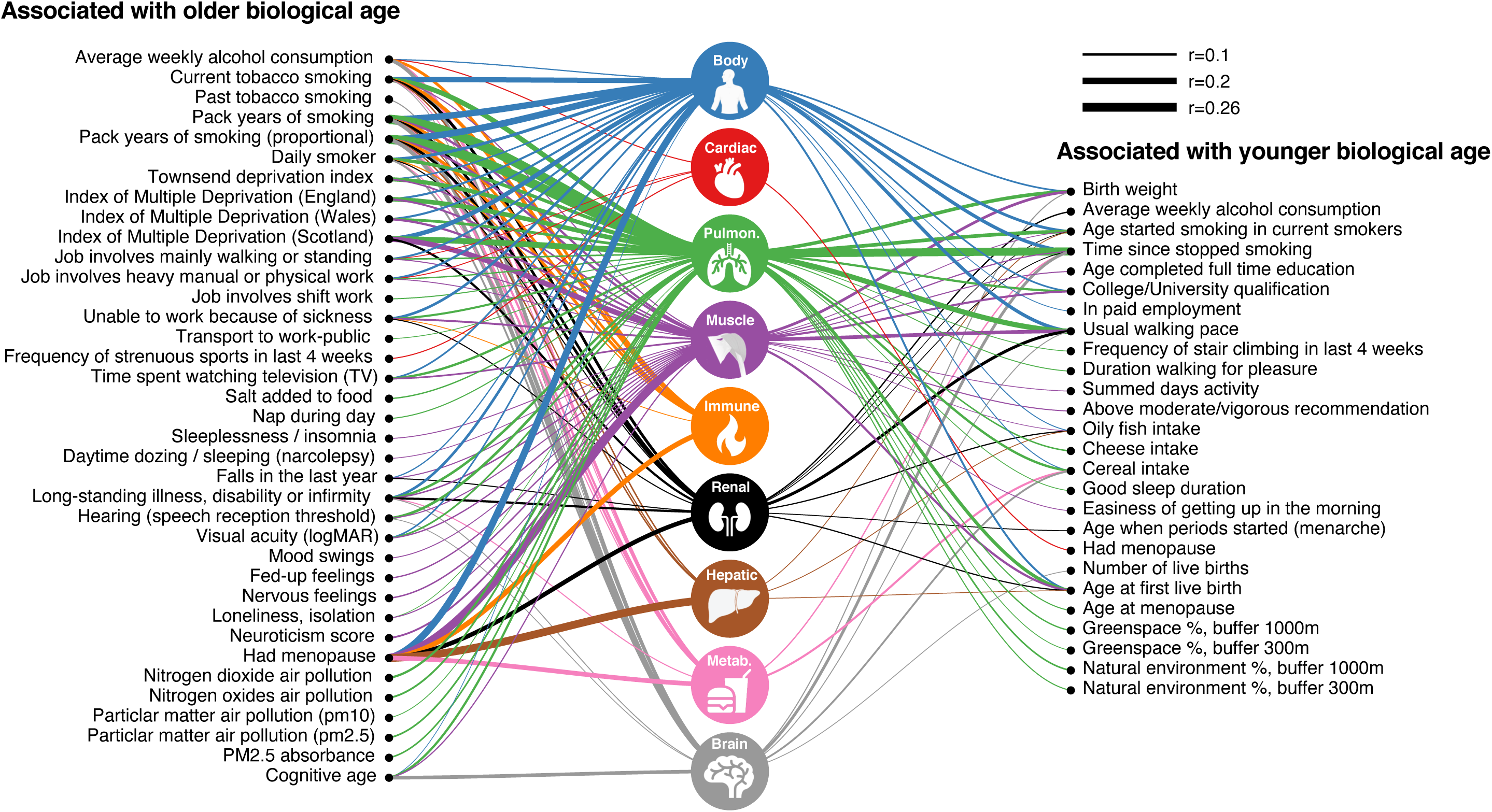
Environmental/lifestyle associations with biological organ age. Icons represent organ systems for which biological age was estimated. Links are shown between environmental/lifestyle factors significantly associated with organ-specific age gaps (p<2.6×10-5, Bonferroni corrected). Links are suppressed for small effect sizes (|r|<0.05). Left (right) list comprises factors positively (negatively) associated with body/brain age gaps. Partial correlation was used to test for associations between organ-specific age gaps and environmental/lifestyle factors, adjusting for chronological age and sex. Link widths are proportional to absolute value of correlation coefficient magnitudes. Environmental and lifestyle factors were assessed using 158 scales tapping early life experience, socio-demographics, lifestyle, psychosocial, local environmental exposure, general health and cognitive age. Results for gray matter, white matter and functional connectivity are shown in fig. S5.

### Biological organ age and chronic disease

To investigate the relationship between biological age and chronic disease risk, individuals with a lifetime diagnosis of a chronic disease were grouped into 16 disease categories: Parkinsonism, multiple sclerosis, stroke, dementia, depression, bipolar disorder, schizophrenia, ischemic heart disease, hypertensive diseases, chronic obstructive pulmonary disease (COPD), chronic kidney disease (CKD), diabetes, cirrhosis, osteoarthritis, osteoporosis and cancer. Additional datasets independent of the UK Biobank were sourced to establish mild cognitive impairment (MCI) and validation dementia cohorts. Disease categories were selected based on lifelong contribution to brain-associated illness burden (i.e., depression, bipolar disorder and schizophrenia), or significant health burden in older adults, including disability and premature mortality^27^. Using the preceding normative models established for healthy individuals, biological age was estimated for each body/brain system and disease category.

Body and brain systems of individuals with chronic disease are significantly older on average than same-aged healthy peers (body: 0.71-6.15 years older; brain: 0.68-4.64 years older). Individuals with CKD have the oldest body ages (mean age gap=6.15±9.32 years) of all 16 disease categories, whereas Parkinsonism associates with advanced body age the least (mean age gap=0.71±5.04 years). Marked heterogeneity in organ-specific ages is evident between and within diseases. Figure 4 shows organs with mean age gaps significantly different from zero in each disease group (p<2.6×10^-4^, Bonferroni corrected for 16 diseases × 12 organs = 192 tests). Organs primarily affected by disease pathology generally show the largest ages gaps on average and show the largest effect sizes (fig. S6). For example, renal, pulmonary, metabolic and hepatic systems are the oldest in CKD (8.03±11.73 years, Cohen’s *d*=0.92), COPD (6.19±6.03 years, Cohen’s *d*=1.26), diabetes (5.17±7.34 years, Cohen’s *d*=0.91) and cirrhosis (4.29 ± 10.21 years, Cohen’s *d*=0.57), respectively. Notably, for many disease categories, organs not typically implicated with disease-specific processes also show evidence of advanced biological age. For example, whereas advanced brain age is evident for most major brain disorders, such as multiple sclerosis (4.64±5.39 years, Cohen’s *d*=1.06), dementia (3.52±5.19 years, Cohen’s *d*=0.75) and Parkinsonism (2.26±4.54 years, Cohen’s *d*=0.58), individuals with non-brain disorders such as diabetes (2.14 ±3.60 years, Cohen’s *d*=0.65), CKD (1.66±3.56 years, Cohen’s *d*=0.50) and COPD (1.65±3.86 years, Cohen’s *d*=0.48) also show significantly advanced brain age (Fig. 4, fig. S5) with moderate effect sizes (fig. S6). CKD-related advanced body and renal ages were replicated in a subgroup of individuals (n=2168, body: 5.85±8.84 years; renal: 7.62 ± 11.08 years) who had not progressed to the end-stage renal disease. Dementia-related advanced brain age was replicated using two additional cohorts (n=284, 3.19 ± 6.13 years, t=16.94, p<2.23×10^-308^). Brain aging was less pronounced in MCI (n=780, 1.07±4.25 years) than dementia (t=10.39, p=3.56×10^-25^), but significantly greater than same-aged healthy peers (t=10.39, p=3.56×10^-25^, fig. S2C). Although rare, some body systems are marginally younger than their chronological age for specific disease categories, including the cardiovascular (schizophrenia), hepatic (diabetes, hypertensive diseases, osteoporosis) and metabolic (osteoporosis, Parkinsonism) systems (Fig. 4B). Disease comorbidity does not explain heterogeneity in organ-specific age across brain versus non-brain disease categories (fig. S7).

**Fig. 4.**
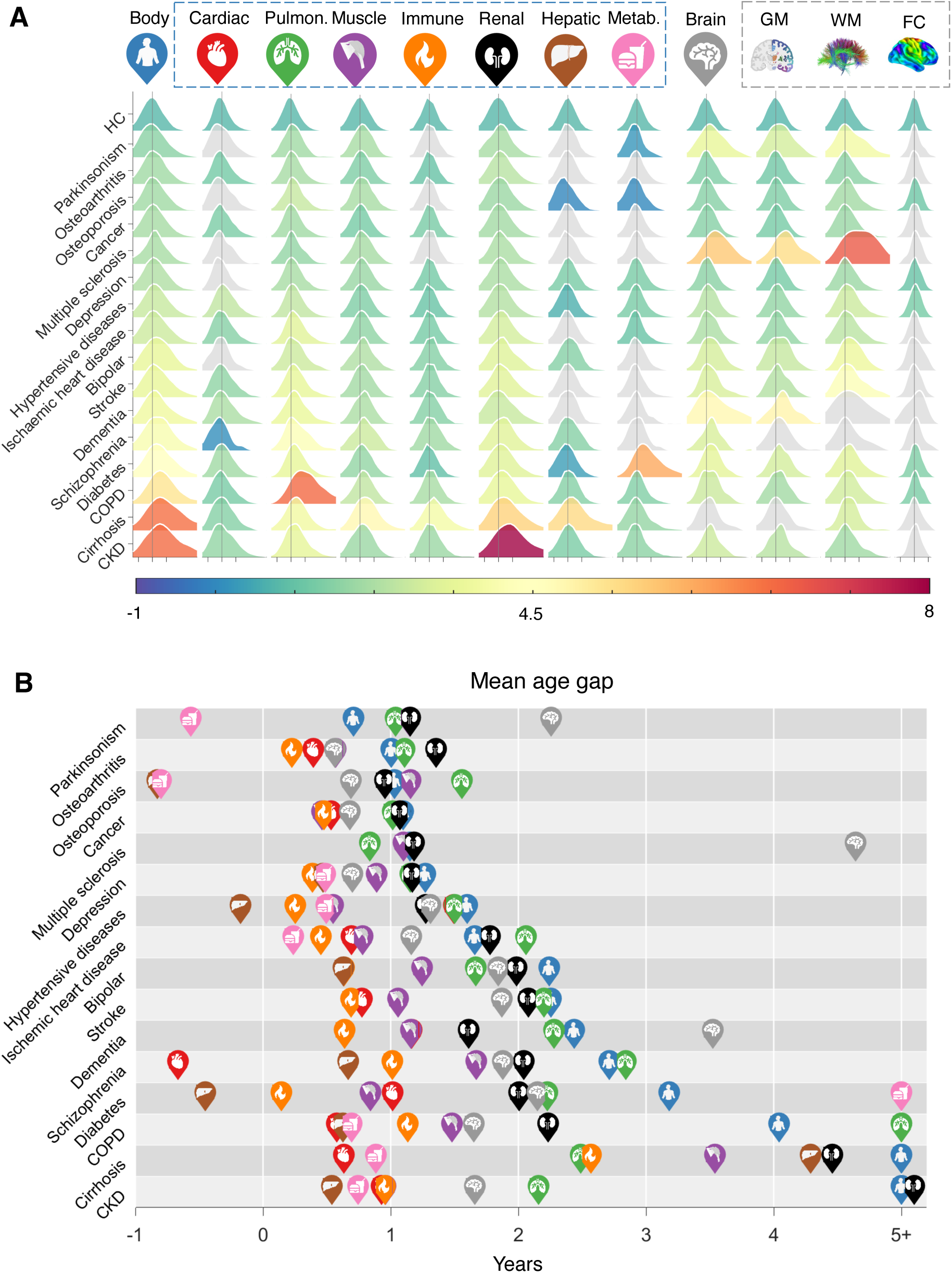
Body and brain age in chronic disease. (**A**) Distribution of body and brain age gaps (columns) for 16 disease categories (rows), compared to healthy individuals (HC). Distributions are colored according to disease- and organ-specific mean age gaps. Colored distributions have a mean that significantly differs from the healthy group (p<2.6 × 10-4, Bonferroni corrected). Despite significant between-group differences, considerable overlap in distributions between disease categories and healthy individuals suggests that factors other than diagnostic status manifest significant heterogeneity of biological age among individuals comprising the same category. Distributions colored gray have a mean that is not significantly different from the healthy group. The three axis ticks on the horizontal axis from left to right for each distribution correspond to age gaps of −5, 0 and 5 years. (**B**) Icons representing body systems and organs are positioned to indicate the mean age gap for each disease category. Icons are not shown for organs with age gaps that do not significantly differ from zero. Organs with age gaps exceeding 5 years are truncated to 5 years for visualization purposes. Disease categories are ordered from top to bottom according to increasing body age gap. COPD, chronic obstructive pulmonary disease; CKD, chronic kidney disease. Metab., metabolic; Pulmon., pulmonary; Muscle, musculoskeletal; Cardiac, cardiovascular; GM, gray matter; WM, white matter; FC, functional connectivity.

We tested whether diagnostic markers confound the interpretation of disease-related aging effects. The exclusion of diagnostic markers for diabetes (i.e., HbA1c) and CKD (i.e., cystatin C and creatinine) from the metabolic and renal aging models, respectively, did lead to decreases in prediction accuracy of chronological age. However, significantly advanced age of the metabolic and renal systems in diabetes and CKD remained evident after these exclusions (see Methods).

Given that some hallmarks of biological aging are also pathological features of aging-related diseases^8^, we hypothesized that phenotypic variation related to chronological age would covary with disease-related phenotypic variation. Consistent with this hypothesis, estimated feature weights of the predictive models of chronological age (one weight per phenotype; number of phenotypes, body: n=78, brain: n=2,309) significantly associate with disease-related phenotypic variation (body: female/male, r=0.43/0.39; brain: female/male, r=0.47/0.52, p<0.0001, Fig. 5A, stratified by disease: fig. S8). Of note, disease and aging phenotypes most strongly associate between body age and osteoporosis-related body changes (female/male, r=0.62/0.48, p<0.003, Bonferroni corrected for 16 disease groups); brain age and ischemic heart disease-related brain changes (female/male, r=0.49/0.55, p<0.003). In contrast, brain age only weakly associates with schizophrenia-related brain changes (female/male, r=0.15/0.15, p<0.003), while diabetes-related body changes do not significantly associate with body age (female/male, r=0.24/0.22, p=0.037/0.056).

**Fig. 5.**
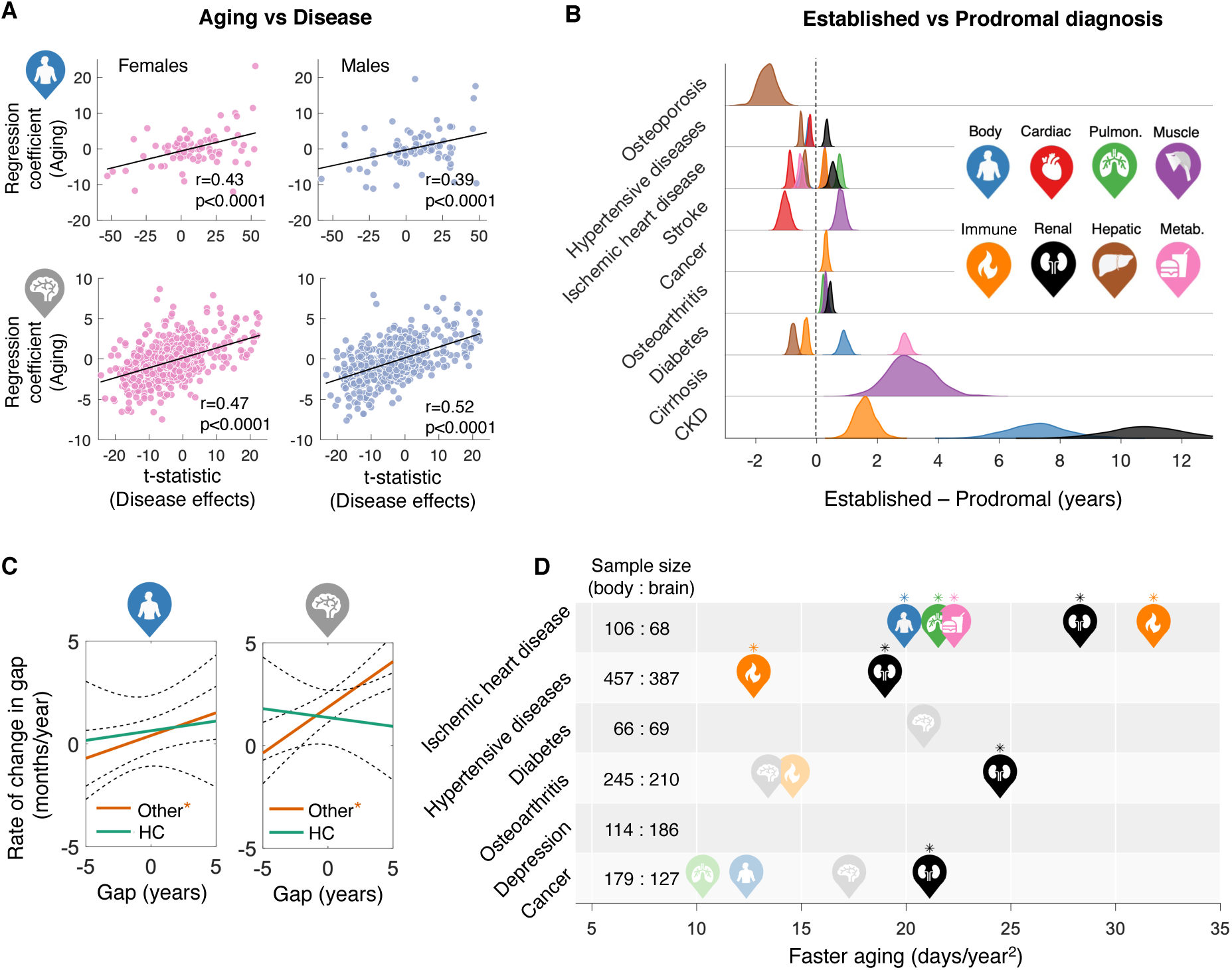
Associations between aging and disease effects and progression. (**A**) Scatter plots show associations between regression coefficients (feature weights) of the age prediction models (y-axis) and between-group differences (disease vs health, t-statistics) in body (n=78, upper) and brain (n=2,309, lower) features (x-axis). The t-statistic quantifies disease-related variation for each phenotype. Each data point represents a phenotype. Lines of best fit indicated with solid black lines. r: Pearson correlation coefficient. Refer to fig. S8 for stratification by disease category. (**B**) Distributions show the difference in age gaps between individuals with established diagnoses at the time of baseline assessment and prodromal individuals who were first diagnosed post assessment, both normalized to age-matched peers. Distributions only shown for organs and disease categories with a significant difference in age gap (two-sample t-test, p<3.9×10-4, Bonferroni corrected). Distributions of the mean differences in age gap were estimated with bootstrapping (n=1000). (**C**) Evidence of faster body (left) and brain (right) aging in chronic disease. Lines of best fit show associations between the rate of change in age gap (y-axis) and the average age gap across the two visits (x-axis). Associations were tested separately in healthy individuals (HC, green) and individuals with chronic diseases (Other, orange). Asterisks indicate significant associations (p<0.0125, Bonferroni corrected). (**D**) Same as panel (C) but stratified according to disease category and organ. Icons are positioned to indicate faster aging, quantified by the slope of the line of best fit between rate of change in age gap and average age gap across the two visits. Associations were only tested for disease categories and/or organs comprising greater than 50 individuals. Icons are shown for organs with a nominally significant association (p<0.05, uncorrected), with high icon opacity and asterisks indicating associations surviving FDR correction of 5%.

### Biological organ age relates to disease progression

For each disease category, we divided individuals into prodromal and established disease groups, based on the date of first diagnosis (if known) and the date of baseline assessment of body and brain function. Individuals who did not experience disease onset/diagnosis before the time of baseline assessment were considered prodromal. Several organ systems of the prodromal groups are significantly older than same-aged healthy peers (fig. S9A), although mean age gaps are larger for individuals with established diagnoses, compared to prodromal individuals (fig. S9B). Hence, advanced body age predates disease diagnosis. Furthermore, between-group differences in mean age gaps (prodromal vs established diagnoses) for several body systems and diseases are significantly larger in groups with established diagnoses, compared to prodromal groups (p<2.6×10^-4^, Bonferroni corrected for 12 organs × 16 diseases = 192 tests), although effect sizes are modest (Fig. 5B). These effects are greatest in CKD (renal: 10.8 ± 1.2 years older), cirrhosis (musculoskeletal: 3.1 ± 0.8 years) and diabetes (metabolic: 2.9 ± 0.1 years), where body systems are significantly older in established compared to prodromal groups.

The rate of change in age gaps significantly associates with the average age gap over the two assessment time points in individuals with chronic disease (body: 𝛽=0.21, p=0.01; brain: 𝛽=0.44, p=4.4×10^-6^; Fig. 5C). This suggests disease-related faster body and brain aging, where each one-year increase in the mean body (brain) age gap associates with a 0.21 (0.44) months/year increase in the rate of body (brain) aging. On the contrary, the rate of aging is constant in healthy individuals (body: 𝛽=0.07, p=0.74; brain: 𝛽=-0.08, p=0.66). Faster aging is evident for multiple organ systems in ischemic heart disease, hypertensive diseases, diabetes, osteoarthritis and cancer (p<0.05, FDR corrected across 6 disease groups × 12 organ systems = 72 tests). Depression shows no evidence of faster aging (p>0.05, Fig. 5D).

### Biological organ age predicts mortality risk

We sought to predict risk of mortality using body and brain age gaps. Mortality was determined using data linkages to national death registries in the UK. Cancer (29.6%), circulatory (26.7%) and respiratory (11.8%) diseases were the three main causes of death (fig. S10). Survival after baseline assessment was ascertained up to 13.41 years for body (n=8,109, age of death 42-83 years, mean 69.6±7.3, 5,670 males) and 6.07 years for brain (n=330, age of death 53-82 years, mean 70.7±6.4, 203 males). Body (2.95±6.56 vs 0.57±4.44 years, p<0.0056, Bonferroni corrected for 9 body ages), and brain (1.72±4.07 vs 0.48±3.31 years, p<0.0125, Bonferroni corrected for 4 brain ages) age gaps significantly differ between deceased and non-deceased individuals. Between-group differences are also evident for specific body (Fig. 6A) and brain systems (fig. S11A).

**Fig. 6.**
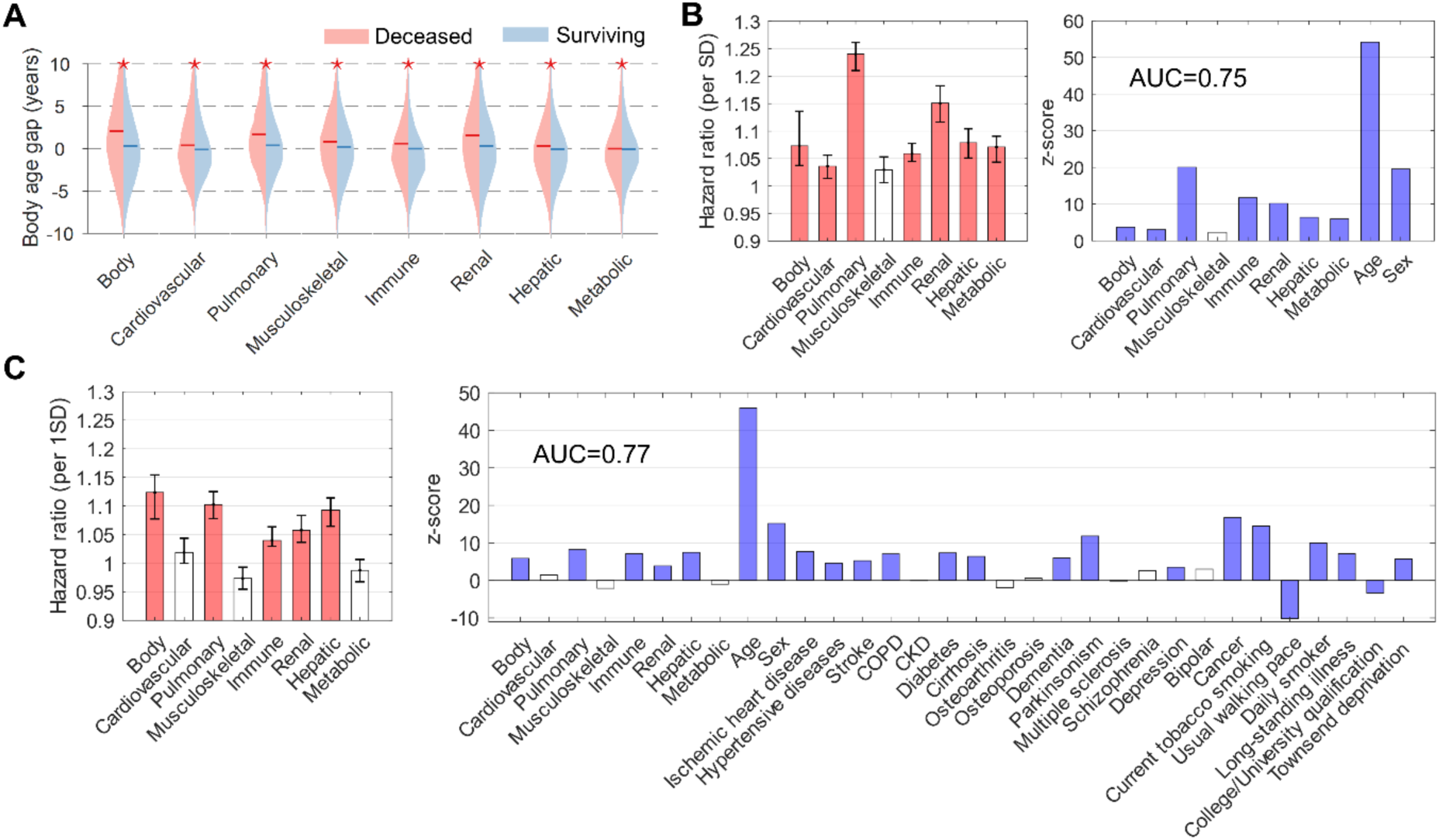
Body age and the risk of mortality. (**A**) Body age gaps in deceased (n=8,109) compared to non-deceased (n=135,314) individuals. Asterisks indicate significant between- group differences, controlling for chronological age and sex (p<0.0056, Bonferroni corrected). (**B**) Bar plots show mortality hazard ratios per one standard deviation (SD) increase in organ- specific age (left) and corresponding z-scores (right). Chronological age and sex are included in the regression. Confidence intervals (95%) estimated with bootstrapping (n=100). Colored bars indicate organs with significant hazard ratios (p<0.005, Bonferroni corrected for 10 dependent variables). AUC: area under curve. (**C**) Same as panel (B), but existing disease diagnoses, general health and key lifestyle factors included in the regression. Colored bars indicate organs with significant hazard ratios (p<0.0015, Bonferroni corrected for 32 dependent variables).

Cox proportional hazards regression, where survival durations were right censored for non-deceased individuals (body: n=135,314; brain: n=36,571), reveal that body age (Fig. 6B,C, table S9), but not brain age (fig. S11B,C, table S10) is a significant predictor of mortality. In particular, adjusting for chronological age and sex, each one standard deviation (SD) increase in a person’s organ age associates with a 7.3% (body, hazard ratio (HR)=1.073, 95% confidence interval (CI)=[1.037, 1.136], p=2.2×10^-6^), 3.6% (cardiovascular, HR=1.036, 95% CI=[1.014, 1.056], p=2.7 × 10^-3^), 24.0% (pulmonary, HR=1.24, 95% CI=[1.210, 1.262, p=2.6×10^-89^), 5.9% (immune, HR=1.059, 95% CI=[1.045, 1.078, p=1.0×10^-31^), 15.1% (renal, HR=1.151, 95% CI=[1.116, 1.183, p=3.0×10^-24^), 7.9% (hepatic, HR=1.079, 95% CI=[1.051, 1.104], p=1.9×10^-10^) and 7.1% (metabolic, HR=1.071, 95% CI=[1.043, 1.091], p=3.2×10^-9^) relative increase in the risk of mortality (area under the curve (AUC)=0.75, loglikelihood=-9.19 × 10^4^, Fig. 6B). This model significantly outperformed a baseline mortality model including only chronological age and sex (AUC=0.72, loglikelihood=-9.29×10^4^, 𝜒^2^=1.86×10^3^, p<2.23 × 10^-308^). Several body systems (i.e., pulmonary, immune, renal, hepatic) remain significant mortality predictors, when controlling for existing disease diagnoses (fig. S12). A regression model including chronological age, sex, all 8 body age gaps, existing disease diagnoses, general health (i.e., long-standing illness and disability) and key environmental/lifestyle factors such as smoking, exercise, tertiary education and socioeconomic inequality yielded the most accurate (AUC=0.771) and best fitting model of mortality risk (loglikelihood=-8.97 × 10^4^, 𝜒^2^=4.98 × 10^3^, p<2.23 × 10^-308^, Fig. 6C). After controlling for the above covariates, the composite body age gap outperformed all organ-specific age gaps in explaining mortality hazard, suggesting that these factors are significant confounds. Replacing body age gaps with body phenotypes associated with mortality, including systolic blood pressure^28, 29^, forced expiration volume in 1-second^30, 31^, hand grip strength^32^, C-reactive protein^33, 34^, serum creatinine^35^, serum alanine and asparate aminotransferase^36^ and the total/high-density lipoprotein cholesterol ratio^37^ did not improve the model accuracy (AUC=0.770) and fit (loglikelihood=-8.98×10^4^, table S11). Similarly, replacing brain age gaps with several global brain measures, including whole brain volume of gray matter, cerebrospinal fluid, white matter, white matter hyperintensity load and mean cortical thickness, mean fractional anisotropy and mean diffusivity did not improve the model (AUC=0.722, loglikelihood=-3.14×10^3^, table S12). Mortality risk associated with body (fig. S13) and brain (fig. S14) age remained largely unchanged after excluding deaths subsequent to the date of coronavirus disease (COVID-19) emergence in the UK^38^ (n=1,033 for body age and n=127 for brain age analyses), suggesting that COVID-19 did not significantly confound our mortality risk estimation models. Further analyses of mortality due to specific disease cause, including cancer (AUC=0.75, fig. S15A), circulatory diseases (AUC=0.84, fig. S15B) and respiratory diseases (AUC=0.86, fig. S15C) reveal similar results, where pulmonary, immune and renal age remain significant predictors for all three causes of mortality (table S13). Finally, logistic models were developed to predict survival time (5-year: AUC=0.774±0.006; 10-year: AUC=0.770±0.003) and premature mortality (death before 70 years old: AUC=0.86±0.003; 75 years old: AUC=0.86±0.003) using body age gaps (fig. S16).

## Discussion

By establishing normative models of aging-related decline for multiple brain and body systems in the world’s largest population-based biobank, we showed that aging is a complex, multisystem process, whereby the biological age of one organ system selectively influences the aging of multiple other systems via characteristic aging pathways. While biological aging is an established concept^12–14^, and earlier studies establish aging clocks for individual organs, including the kidneys^39^, heart^40^, lungs^41^, skin and blood^42^, we derived the first whole-body multiorgan characterization of aging. Our organ clocks enabled elucidation of unique organ age profiles for 16 chronic diseases and discovery of modifiable factors that can potentially lead to disease-specific longevity interventions targeted at specific body systems, ultimately extending lifespan.

Our work enhances the clinical utility of proxy measures of aging developed for older individuals, such as frailty indices^43^, as well as existing DNA methylation (epigenetic) clocks^44, 45^. While epigenetic clocks are clinically useful and provide important insights into aging biology across tissue types, it is now recognized that aging varies markedly between organ systems and tissues, particularly in disease^46^. Bespoke organ and disease-specific aging clocks are thus needed to enhance the clinical utility of existing pan-tissue clocks, which do not readily differentiate between tissue components and body systems^15^. Addressing this need, we showed that deviations from normal aging-related decline can be detected in certain organs (but not all) years before disease diagnosis. These deviations predict mortality, even after controlling for chronological age, disease burden and other risk factors. Our organ clocks could thus be used to identify individuals in midlife, before disease onset, who may benefit from early interventions aimed at slowing the aging of specific body systems and organs.

Crucially, as with the frailty indices^43^, many of the biological markers that inform our organ clocks are already widely assayed in primary care (e.g., full blood counts, renal and liver function, blood pressure, lipids, glucose), are readily accessible at minimal cost (forced respiration, grip strength, waist circumference), or are accessible and cost effective when benchmarked against the burden of chronic illness (brain MRI scans). Alongside the relatively modest computational burden of the model algorithms (especially when pretrained), these considerations argue for direct, cost effective and feasible clinical implementation of organ age in primary care.

Our investigation into environmental and lifestyle factors can inform real-world personalized interventions targeted at specific body systems, through change of lifestyle, such as limiting tobacco smoking and alcohol intake, exercise, education, sleep hygiene and maternal nutrition, as well as efforts requiring national inputs such as reductions in socioeconomic inequality and air pollution, and improvements in residential greenspace and natural environment coverage. Studying the impact of such interventions would provide causal evidence for the conditional effects presently reported. The pulmonary, metabolic and immune systems are promising organ-specific targets for interventions, given that these systems influence the rate of aging of multiple body systems (i.e., cardiovascular, musculoskeletal), via interorgan aging pathways (Fig. 2E). Notably, the aging pathway linking pulmonary, cardiovascular and musculoskeletal systems recapitulates the known epidemiological link between impaired lung function, weak muscle strength and elevated risk of adverse cardiovascular outcomes^30, 32^. While aging-related brain gray matter loss is normal^47^, we found that advanced gray matter age substantially influences the rate of aging-related decline in brain connectivity, but not the converse (Fig. 2F). Given that body phenotypes were measured several years before brain phenotypes (4-14 years earlier; Fig. 2C), the estimated influence of advanced body age, particularly of the cardiovascular system on brain age (Fig. 2D), may thus reveal early signs of brain aging.

While the organs that manifest primary disease processes appear the oldest in individuals with the disease, we found that advanced organ age is widespread, involving multiple body and brain systems. Brain systems of individuals with non-brain disorders, including diabetes, chronic kidney, pulmonary and cardiovascular diseases appear significantly older than same-aged healthy peers, whereas body systems, particularly pulmonary and renal systems, show signs of advanced aging in individuals primarily diagnosed with major brain disorders, including schizophrenia, dementia, bipolar disorder, depression, multiple sclerosis and Parkinsonism. It is important to acknowledge that some of our clocks are informed by diagnostic markers, which may potentially confound the interpretation of disease-related differences in biological age. However, confounding can be ruled out for the above examples where advanced age is evident for organs that are not informed by diagnostic markers relevant to the disease under consideration. Furthermore, excluding key diagnostic markers reduced model performance but did not alter our conclusions about the association between disease and organ aging.

Chronological age and male sex were found to be the two strongest mortality risk factors, consistent with previous literature^48, 49^. After controlling for these two factors, organ ages remained strong mortality risk factors, particularly pulmonary age, followed by ages of the renal, hepatic, metabolic, immune and cardiovascular systems. Individuals who subsequently deceased had older appearing brains compared to those who survived, consistent with a previous study examining brain age and mortality^50^. However, advanced brain age did not predict increased risk of mortality. This may be due to the relatively low mortality rate in individuals with brain age estimates in the UK Biobank (mortality rate: 330/36901=0.0089) compared to the Lothian Birth Cohort 1936 used by Cole and colleagues (mortality rate: 73/669=0.11). Continued follow-up of UK Biobank participants will likely yield more insight into the relationship between brain aging and mortality.

Advanced pulmonary age was the strongest predictor of mortality (HR: 1.24), consistent with epidemiological observations of associations between impaired lung function and increased risk of mortality^30, 31^. While reduced muscle strength, measured by handgrip strength, is commonly associated with increased risk of mortality^51^, older musculoskeletal age was not a significant risk factor of mortality when controlling for chronological age, sex and the age gaps of other organs. This is consistent with the configuration of the multiorgan aging network, where the musculoskeletal system is a central hub, influenced by the extent of aging of most other organ systems. The mortality risk explained by musculoskeletal aging may thus be attributable to the biological age of other body systems. Advanced age of the pulmonary, immune, renal and hepatic systems significantly raises a person’s mortality risk, beyond that explained by existing chronic diseases, chronological age and sex (Fig. 6). Whereas disease conditions that primarily affect these organ systems are common causes of death^27^, our results demonstrate the uniqueness of biological age in explaining all-cause mortality, regardless of existing diseases.

The divergence of an organ’s biological age from chronological age may emerge early in life and widen over the lifespan, increasing the risk of chronic disease and mortality. However, the *rate* of aging reported here more specifically reflects aging-related decline rather than early life events. Interventions designed to delay the rate of organ aging may thus effectively delay disease onset, resulting in an extended healthy lifespan. Further study is needed to determine whether interventions informed by the observational evidence reported here can reduce these risks and potentially slow organ aging in at-risk individuals. Further work is also needed to determine the genetic influences on our organ clocks. We showed that leucocyte telomere lengths and genetic variants known to index leucocyte telomere length weakly associate with several body ages. This complements a recent genome-wide association study^13^ showing the importance of the immune system (major histocompatibility complex on chromosome 6) and DNA repair pathways in aging.

Our work addresses several recently identified challenges hindering the clinical translation of biological aging research^15, 19^. We established bespoke clocks that measure the biological age of specific brain and body systems using markers that are routinely assayed in primary care, elucidated organ age profiles for prevalent chronic diseases and identified modifiable factors that can inform new strategies to potentially limit organ-specific aging. Our work is the first to map a multiorgan aging network for the human body.

Several caveats pertain to our findings. First, biological aging is multifaceted. As such, it is unlikely that a single index of organ aging will be sufficient and conclusive. As with the continued refinement of epigenetic clocks over the last decade^44, 52–54^, and ongoing deliberations about how to define frailty in older people^43, 55^, standardized measures of organ age remain to be developed. Second, body phenotypes were measured several years before brain phenotypes in the UK Biobank. Due to the sequential and non-randomized participant assessment schedule, we were unable to assess the influence of brain aging on body systems. Future investigations, leveraging multiple cohort waves, may reveal bidirectional brain-body influences. Third, most participants enrolled in the UK Biobank are from a white ethnic background. Inclusion of participants from a diversity of ethnicities, demographic and socioeconomic backgrounds will be required to assess the generalizability of our current finding. Finally, some imaging modalities (e.g., carotid imaging) were only acquired in select individuals, limiting the data available for some organ clocks. Clinical translation could proceed by adding these to clinical assays, and/or removing others based upon the trade-off between their added predictive value versus their cost.

## Methods

### Participants

Individuals (n=502,504, 229,122 males) participating in the UK (United Kingdom) Biobank^56^ were analyzed for the primary study (Project ID 60698). They were aged 37-73 years at the time of recruitment (2006-2010) and underwent extensive questionaries, physical assessments, blood and urine sample assays and genome-wide genotyping at 22 assessment centers throughout the UK. A subset of individuals (n=20,345, 9,938 males) was followed up during 2012-2013 for repeated physical and physiological assessments. Multimodal brain imaging ^57^ was acquired during the third visit (2014-2020) at three mirrored imaging centers located at Manchester, Reading and Newcastle, respectively, in 49,002 individuals (23,710 males). Follow-up brain imaging was conducted from 2019 onwards in 1,503 individuals (754 males), providing a longitudinal sample enabling estimation of the rate of change in biological age. An assessment timeline is shown in Fig. 2C. Each step in the assessment and processing of biological samples was handled and monitored centrally to minimize biases across recruitment centers. We found that biological age (i.e., age gaps) showed negligible site-related (fig. S17), ethnicity-related (fig. S18) and longitudinal subsampling-related (fig. S19) variation. The UK Biobank has approval from the North West Multi-centre Research Ethics Committee (MREC) to obtain and disseminate data and samples from the participants (http://www.ukbiobank.ac.uk/ethics/). Written informed consent was obtained from all participants. Details of participants comprising the independent validation cohorts are described below (see *Validation of brain age prediction using external datasets*).

### Body age phenotypes

Physical and physiological measures known to index the function, structure and/or general health of the cardiovascular, pulmonary, musculoskeletal, immune, renal, hepatic and metabolic systems were selected, resulting in 101 organ-specific phenotypes. Physical measures included standing height, weight, body mass index (BMI), hip and waist circumferences, handgrip strength, ultrasound heel bone densitometry, spirometry and cardio-respiratory fitness. Physiological assessments included blood pressure, pulse rate, arterial stiffness, blood hematology, blood and urine biochemical assays. Further steps included:

1. Averaging measures if tested for left and/or right side of the body. E.g., handgrip strength, heel bone mineral density and ankle spacing width.
2. Averaging measures if tested more than once at the same visit. E.g., diastolic and systolic blood pressure and pulse rate were each measured twice.
3. Selecting the best performance among multiple repeated tests at the same visit. E.g., spirometry test for lung function, including forced vital capacity (FVC), forced expiration volume in 1-second (FEV1) and peak expiratory flow (PEF). Each participant was asked to conduct up to three blows (lasting for at least 6 seconds) within a period of approximately 6 minutes. The quality of each blow result was automatically detected by the device and only the best performing blow of acceptable quality was selected. The FEV1/FVC ratio was also computed and used for body age estimation.
4. Excluding measures with missing responses in more than 30% of individuals. As such, measures of cardio-respiratory fitness (missing proportion: 85%), arterial stiffness (69%), urine microalbumin (76%), blood oestradiol (79%) and Rheumatoid factor (93%) were excluded.

This resulted in 78 body phenotypes for chronological age prediction (table S1). Participants with missing responses for any of the 78 phenotypes were then excluded, resulting in a final sample comprising 143,423 individuals (age range 39-73 years, mean 56.7±8.2 at the baseline assessment of body function, 79,980 males). Follow-up data were available in 1,220 individuals (age range 44-75 years, mean 61.6±7.7 at the second visit, 837 males) at 2.1-5.6 years follow-up. Phenotypes were grouped based on relevance to the structure and function of each organ systems, forming 7 phenotype groups. A predictive model of chronological age was established using all phenotypes comprising a given organ-specific phenotype group (see below). Additionally, a whole-body predictive model was established using all body phenotypes, irrespective of organ grouping. Key measures used to assess individual organ function are as follows (also see Fig. 1A):

- Cardiovascular system: pulse rate, systolic blood pressure and diastolic blood pressure.
- Pulmonary system: FVC, FEV1, PEF and FEV1/FVC ratio.
- Musculoskeletal system: handgrip strength, standing height, weight, BMI, waist and hip circumference, waist/hip circumference ratio, heal bone mineral density, ankle spacing width, blood biochemical markers such as phosphatase, calcium, phosphate and vitamin D.
- Immune system: C-reactive protein and blood hematology tests of leukocytes, erythrocytes, thrombocytes and hemoglobin.
- Renal system: biomarkers associated with glomerular filtration and electrolyte regulation, including creatinine (enzymatic), potassium and sodium in urine, albumin, urea, urate, creatinine, cystatin C, phosphate and total protein in blood.
- Hepatic system: alanine aminotransferase, aspartate aminotransferase, gamma-glutamyl transferase, direct and total bilirubin, albumin, alkaline phosphatase and total protein in blood.
- Metabolic system: blood biomarkers associated with lipids and glucose metabolism, including apolipoprotein A, apolipoprotein B, cholesterol, glucose, glycated hemoglobin, high-density lipoprotein cholesterol, direct low-density lipoprotein cholesterol, Lipoprotein A and triglycerides.

Several blood biomarkers, including insulin-like growth factor 1, testosterone and sex hormone-binding globulin were not assigned to any of the seven systems and were only used for overall body age gap estimation. Post-hoc analysis was performed to investigate the potential confounding effect of antihypertensive medications (i.e., angiotensin-converting enzyme inhibitors, angiotensin receptor blockers, beta-blockers, calcium channel blockers, thiazide diuretic agents) on the estimation of cardiovascular age. This grouping of medication categories is consistent with a previous UK Biobank study^58^. Adjusting for chronological age and sex, we found no significant difference in the estimated cardiovascular age between individuals who regularly take antihypertensive medications (mean age gap=0.32±3.58 years) and individuals who do not take any antihypertensive medications (mean age gap=0.46±3.94 years, t=1.04, p=0.29), suggesting that antihypertensive medications are not significant confounds.

### Brain age phenotypes

Structural and functional brain phenotypes (n=2,309, table S2) derived from 3 neuroimaging modalities, including T1-weighted magnetic resonance imaging (MRI), diffusion MRI (dMRI) and resting-state functional MRI (fMRI) were sourced from the UK Biobank^57, 59^. The image processing pipeline, artefact removal, cross-modality and cross-individual image alignment, quality control and phenotype calculation are described in detail in the central UK Biobank brain imaging documentation (https://biobank.ctsu.ox.ac.uk/showcase/showcase/docs/brain_mri.pdf) and by Alfaro-Almagro and colleagues^59^. Participants with missing entries for any of the 2,309 phenotypes were discarded, resulting in a final sample comprising 36,901 individuals (age range 45-82 years, mean 64.2±7.5 at the first imaging visit, 17,203 males) for brain age analyses. Repeated brain imaging phenotypes were available in 1,294 individuals (age range 50-83 years, mean 65.2±7.2 at the second imaging visit, 632 males) at 2.0-2.7 years follow-up.

Predictive models of chronological age were established using imaging-derived phenotypes (IDPs) pertaining to gray matter structure, white matter microstructure and brain functional connectivity (FC). Additionally, a whole-brain predictive model was established using all brain phenotypes (n=2,309), irrespective of brain tissue class. IDPs used to assess individual brain systems are as follows:

- Gray matter: regional gray matter volume, cortical thickness and surface area, as derived from T1-weighted MRI (number of IDPs: 578).
- White matter: dMRI-derived microstructural measures of white matter tracts including mean fractional anisotropy and mean diffusivity (number of IDPs: 92)
- Brain functional connectivity (FC): connectivity strengths between 55 functional brain networks derived from resting-state fMRI (IDPs: 1,485 connection pairs)

Other IDPs such as regional gray/white matter intensity contrast from T1-weighted MRI and volumes of ventricles were only included in the whole-brain predictive model.

### Cognitive phenotypes

Cognitive tests assessing reasoning, memory, attention, processing speed and executive function were conducted on the same day of brain imaging in 45,930 individuals (22,307 males). A predictive model of chronological age was established using 29 distinct measures of cognitive performance (table S3). Dummy variables were generated for categorical responses. Several cognitive tests, including trail making, matrix pattern completion, tower rearranging and symbol digit substitution test were only added to the assessment battery from 2016 onwards, resulting in an incomplete assessment for some participants. These participants were omitted, yielding a final sample comprising 32,317 individuals (age range 45-82 years, mean 65.1±7.6, 15,712 males) for cognitive age analyses.

### Normative aging models

Support vector machines (SVMs) were trained to predict an individual’s chronological age using body (n=28,589, age range 40-70 years, mean 52.7±7.8, 15,444 males), brain (n=7,922, age range 46-82 years, mean 61.8±7.3, 3624 males) and cognitive (n=7,167, age range 47-82 years, mean 62.6±7.3, 3,357 males) phenotypes in healthy individuals, defined as no self-reported and healthcare documented lifetime chronic medical conditions (see Health outcomes and clinical characterization). Compared to linear regression, SVM regression can provide improved robustness to outliers and overfitting. It automatically learns the relative value of each phenotype toward predicting age and fits a hyperplane to the phenotype data. Using 20-fold cross-validation, predictive models were developed for each body (n=8) and brain (n=4) system as well as for cognitive performance (Fig. 2B). Separate models were trained for males and females. Each model accepted an individual’s organ-specific phenotypes and yielded an estimate of chronological age based on these phenotype inputs. Individuals were thus characterized by 12 organ-specific predictions of chronological age.

For each 20-fold cross-validation iteration, a linear SVM was trained to predict chronological age using individuals comprising 19 folds (training set). The fitted regression coefficients (feature weights) were then applied iteratively to the held out set of individuals (test set), resulting in a predicted chronological age for each healthy individual. In this way, the prediction model was never trained using the same individuals for which it was applied to predict age, minimizing the risk of overfitting. All measures except for categorical variables were standardized by weighted column mean and standard deviation, computed within the training set prior to each iteration of model training. For all models, the SVM box constraint and kernel scale were set to unity, and the half-width of the epsilon-insensitive band was set to a tenth of the standard deviation of the interquartile range of the predicted variable (i.e., chronological age). The SVM was solved using sequential minimal optimization, using a gap tolerance of 0.001. More specifically, linear SVM regression involved fitting the linear function,

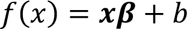

for each organ system, where 𝒙 is the matrix of organ-specific phenotypes (subjects × phenotypes), 𝜷 is the fitted model coefficients and 𝑏 is the model offset. To estimate 𝜷 and 𝑏 for each organ system, the following objective function was minimized,

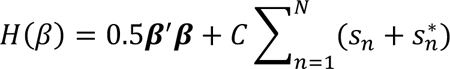

subject to the constraints |𝑎𝑔𝑒_𝑛_ − (𝒙_𝑛_𝜷 + 𝑏)| ≤ 𝜖 ; 𝑎𝑔𝑒_𝑛_ − (𝒙_𝑛_𝜷 + 𝑏) ≤ 𝜖 + 𝑠_𝑛_ ; (𝒙_𝑛_𝜷 + 𝑏) − 𝑎𝑔𝑒_𝑛_ ≤ 𝜖 + 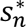 and 𝑠_𝑛_, 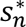 ≥ 0 for all individuals 𝑛 = 1, … 𝑁 in the training data set. In this formulation, 𝑠_𝑛_ and 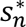 are the slack variables for each individual, 𝜖 is the model residual and 𝐶 is the box constraint constant (𝐶 = 1 in this work). Consistent with recent work^60^, using a non-linear kernel function (i.e., Gaussian or polynomial) did not improve model performance. The predicted chronological age of the individual with index 𝑛 comprising the test dataset was given by

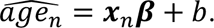

Model performance was quantified using the Pearson correlation coefficient (r) and mean absolute error (MAE) between predicted and chronological age in the test sets. The 20-fold cross-validation procedure was repeated for 10 trials, randomizing the assignment of individuals to folds for each trial. Owing to the large sample size, variation across different train-test data splits was negligible (r_sd_<0.002, MAE_sd_<0.005; sd: standard deviation). Optimization of hyperparameters (i.e., box constraint, kernel function, epsilon) did not substantially improve performance of the predictive models trained on all body or brain phenotypes, and thus hyperparameter optimization was not conducted for the organ-specific models.

To determine chance-level prediction accuracy intervals, chronological age was randomly permuted among individuals and each organ-specific predictive model was re-trained using the permuted data. This was repeated for 5000 permutations to generate an empirical null distribution for MAE, under the null hypothesis of an absence of predictive utility of body and brain phenotypes on chronological age prediction. The observed MAE for all predictive models was less than the 5th percentile of the MAE null distribution, enabling rejection of the null hypothesis. The false discovery rate was controlled at 5% across all predictive models (n=26) using the Benjamini-Hochberg procedure^61^.

Prediction accuracies varied considerably between organ systems (Fig. 2A). While prediction accuracy often improved with the number of features available, this was not always the case. For example, the least accurate organ-specific model (i.e., immune system) comprised the largest number of features (n=33), whereas the best performing model comprised 11 phenotypes pertaining to renal function. Models developed for the pulmonary (n=4) and cardiovascular system (n=3) also outperformed the immune system. Prediction accuracy variation between organ models could be due to i) insufficient or inaccurate phenotype ensembles to fully characterize an organ’s age-related decline; or ii) complex trajectories of age-related decline that are nonlinearly related to chronological age. Regarding the latter consideration, deep neural networks and nonlinear learners could have improved the prediction accuracies reported here, as suggested in recent brain age prediction studies^60, 62–65^. Regarding the former, we note that most physiological measures (e.g., blood biochemistry, urine assays) used in this study are validated with rigorous quality control procedures (https://biobank.ctsu.ox.ac.uk/crystal/ukb/docs/biomarker_issues.pdf) and are commonly used in clinical settings as diagnostic tools to assess organ-specific function and general health. For example, elevated serum liver enzyme levels often reflect hepatocyte damage or cholestasis^66^. However, key phenotypes for some body systems were unavailable. For example, inflammatory cytokines would have enabled a more holistic characterization of immune function^67^, while cardiac and carotid imaging would have enabled detailed assessment of heart function and atherosclerotic plaque morphology^68^. Of note, cardiac and carotid imaging were not primarily used to estimate cardiovascular age in our study as they were available from 2014 onwards (third visit) in the UK Biobank. The lack of temporal correspondence between cardiac and carotid imaging data and other body phenotypes (i.e., blood biochemistry and urine assays) precluded a concurrent investigation of biological age across multiple body systems. In supplementary analyses, we established a revised cardiovascular normative model that includes heart MRI and carotid ultrasound data and compared the accuracy of the revised model to our original cardiovascular model. We found modest improvement in the age prediction using the revised model (12% and 15% reduction in MAE in females and males respectively, fig. S20). This suggests that our original model using blood pressure indices and pulse rate provides a reasonable estimation of cardiovascular age. Recent work^13^ measures cardiovascular age using a combination of blood pressure, blood markers (e.g., glucose, lipids) and physical fitness (e.g., vital capacity). In contrast, we use blood-derived glucose and lipids, and vital capacity measures to instead inform aging models of the metabolic and pulmonary system, respectively. This exemplifies the need for future standardization of organ age measures, and we suggest that cardiovascular age, as measured by Nie and colleagues^13^, is based on a broader characterization of the cardiovascular system and combines some features of our metabolic and pulmonary age models. In supplementary analyses, we also established revised metabolic and renal normative models that exclude diagnostic markers for diabetes (i.e., HbA1c) and CKD (i.e., cystatin C and creatinine), respectively. We found that prediction accuracy worsens (without HbA1c: females: r=0.45, MAE=5.48; males: r=0.19, MAE=6.59) compared to our original model (with HbA1c: females: r=0.50, MAE=5.30; males: r=0.23, MAE=6.51). However, the metabolic system still shows significantly advanced biological age in diabetes (age gap=0.18±4.64 years, t=6.02, p=1.79 × 10^-9^) after excluding this diagnostic marker. Similarly, a model excluding cystatin C and creatinine led to less accurate prediction of chronological age (females: r=0.43, MAE=5.61; males: r=0.36; MAE=6.15) compared to our original model (females: r=0.53, MAE=5.22; males: r=0.45; MAE=5.83). Nevertheless, the renal system still shows significantly advanced age in CKD (age gap=3.14±6.32 years, t=35.52, p<2.23×10^-308^) after excluding these two diagnostic markers.

Age prediction models trained using healthy individuals were applied to predict the chronological age of individuals diagnosed with one or more diseases (see Health outcomes and clinical characterization). For this purpose, all models were re-trained on the full sample of healthy individuals.

### Age gap index

Subtracting actual chronological age from predicted chronological age, referred to as the *age gap*, provides a normalized measure of the extent to which an individual’s organ system appeared older (gap>0) or younger (gap<0) than same-aged peers of the same sex. Age gaps were estimated for each organ, yielding a multiorgan assay of biological age for each individual. Chronological age was regressed from all estimated age gaps to adjust for regression-toward-the-mean bias^69, 70^ and the residuals of this regression defined adjusted age gaps. All age gaps in this study were adjusted as such. Regressing the square of chronological age in addition to chronological age had minimal impact on the adjusted age gaps. Regression coefficients for performing age gap adjustment were fitted using the training set and then used to adjust the age gaps for individuals comprising the test set. Legacy studies^71–75^ typically quantify biological age using a linear combination of chronological age and selected physiological phenotypes^13, 76–78^, and are sometimes referred to as “Phenotypic Age”^12, 14, 53, 79^. In contrast, for the age gap index used here, chronological age is the prediction target (i.e., independent variable). An advantage of the age gap index is that it is an inherently personalized measure, cross-validated, independent of chronological age, and thereby directly indexes deviations from population norms. Likewise, compared to the commonly used frailty index^43^, which characterizes an overall functional decline in older people (usually > 65 years) by counting the number of health deficits present^80^, an advantage of the age gap index is that it is an organ-specific aging measure applicable across the lifespan.

Longitudinal assessments enabled estimation of the rate of change in age gaps, providing organ-specific estimates of the rate of aging^3, 14^. Of note, all normative models were trained using phenotypes measured at baseline and subsequently applied to predict chronological age in the follow-up data. Let *gap*_t0_ (years) be the organ age gap estimated at baseline, *gap*_t1_ (years) be the age gap at follow-up and *T* be the time interval (in years) between baseline and follow-up assessment. The rate of aging was estimated as Δ = 12×(*gap*_t1_−*gap*_t0_)/*T*, expressed in units of months/year. To test for faster organ aging, the rate of change, Δ, was regressed against the average age gap, (*gap*_t1_ + *gap*_t0_)/2, and a significant positive association between these quantities across individuals provided evidence consistent with faster aging^26^. The slope of the regression line provided an estimate of the putative acceleration rate (months/year^2^). Note that this is effectively a population-level estimate, and it does not necessarily imply faster or accelerated aging for any individual participant.

### Replication of brain age prediction in additional datasets

To assess our normative aging model in an older age cohort, we performed supplementary analyses combining brain MRI data from the Australian Imaging, Biomarkers and Lifestyle Flagship Study of Ageing (AIBL, n=650) (https://aibl.csiro.au/) and the Alzheimer’s Disease NeuroImaging Initiative (ADNI, n=1,677) (http://adni.loni.usc.edu/). The two cohorts comprise individuals diagnosed with mild cognitive impairment (MCI) and dementia as well as healthy individuals, thus facilitating external validation for our normative brain aging model and the relationship between brain age and neurodegenerative diseases.

AIBL study methodology has been reported previously^81^. The AIBL study was approved by the institutional ethics committees of Austin Health, St Vincent’s Health, Hollywood Private Hospital and Edith Cowan University, and all volunteers gave written informed consent before participating in the study. ADNI was launched in 2003 as a public-private partnership, led by Principal Investigator Michael W. Weiner, MD. The primary goal of ADNI has been to test whether serial magnetic resonance imaging (MRI), positron emission tomography (PET), other biological markers, and clinical and neuropsychological assessment can be combined to measure the progression of mild cognitive impairment and early Alzheimer’s disease. For up-to-date information, see www.adni-info.org. As per ADNI protocols, all procedures performed in ADNI studies involving human participants were in accordance with the ethical standards of the institutional and/or national research committee and with the 1964 Helsinki declaration and its later amendments or comparable ethical standards. More details can be found at adni.loni.usc.edu.

Details of brain image acquisition can be found elsewhere^81, 82^. T1-weighted MRI brain images acquired at baseline assessments were used in this study. Consistent with the brain image processing pipeline used for the primary cohort (UK Biobank), MRI brain images were processed using FreeSurfer v6^83^, resulting in 578 regionally specific MRI-derived phenotypes representing regional gray matter volume, cortical thickness and surface area. The Destrieux atlas^84^ was used for cortical parcellation. Additional segmentations of hippocampal subfields^85^, amygdala^86^ and thalamic^87^ nuclei and brainstem substructures^88^ were performed using FreeSurfer v7. The quality of the T1 images was automatically assessed using the Euler number, an index generated by FreeSurfer that measures the topological complexity of a reconstructed cortical surface^89^. Following previous recommendations^90^, images with a Euler number less than -217 were associated with poor quality and thus discarded (AIBL: n=111; ADNI: n=104). Images with any MRI-derived phenotypes residing more than six standard deviations from the median were also discarded (AIBL: n=7; ADNI: n=17).

Given the older age range of the AIBL and ADNI cohorts compared to the UK Biobank, the normative aging model established in the UK Biobank cohort could not be directly applied to the two external datasets. We therefore re-trained the brain age prediction model for gray matter phenotypes in a combined group of healthy individuals across the three datasets. Before model training, data harmonization was performed using ComBat (https://github.com/Jfortin1/ComBatHarmonization)^91, 92^ to control for variation in brain phenotypes due to differences in scanners and datasets. Of note, images acquired from MRI scanners with less than 10 scanned individuals were further discarded (n=186, ADNI) to ensure reliable harmonization, resulting in 532 AIBL (age range 55-96 years, mean 72.4±6.4, 218 males, 4 scanners) and 1,370 ADNI (age range 50-95 years, mean 72.7±7.5, 656 males, 76 scanners) individuals for further analyses. Age, sex and diagnostic status were included as biological covariates in the harmonization.

Consistent with our main findings in the UK Biobank cohort, we found that chronological age could be predicted with high accuracy using gray matter phenotypes (female: r=0.82, MAE=3.52; male: r=0.85, MAE=3.41, fig. S2A). Gray matter feature weights were highly consistent between the original and the re-trained model (female: r=0.86, p<2.23×10^-308^; male: r=0.87, p<2.23×10^-308^, fig. S2B). The re-trained brain age prediction model was applied to estimate brain gray-matter age for individuals diagnosed MCI and dementia. We found that brain age appears significantly older in individuals diagnosed with MCI (n=780, mean age gap=1.07±4.25 years, p=3.56×10^-25^) and dementia (n=284, mean age gap=3.19±6.13 years, p<2.23×10^-308^) than same-aged healthy peers (fig. S2C).

### Structural equation modeling

Structural equation modeling was used to infer the influence of each organ’s baseline age gap on the follow-up age gap (Fig. 2D), or rate of aging, Δ, (Fig. 2E,F) of other organ systems. The fast-greedy equivalence search (FGES) heuristic for continuous variables was performed to search for causal Bayesian networks and determine the highest scoring model. FGES is a Bayesian heuristic that starts with an empty graph and adds edges to improve the score function (i.e., Bayesian Information Criterion, BIC), until no more edges can be added. It then performs a backward search that removes edges, until no edge removal increases the score function ^93^. The search was constrained to edges modeling influences consistent with the flow of time. To this end, age gaps measured at the same assessment were forbidden from influencing each other, age gaps measured at follow-up were forbidden from influencing age gaps measured at baseline, and rates of aging were forbidden from influencing baseline age gaps. The FGES heuristic was repeated for 500 bootstrapped samples and edges present in 50% of the samples formed a final consensus network structure. Regression was used to estimate residual variances and coefficients (𝛽) for the edges comprising the final consensus network. Sex, age at baseline and whole-body (Fig. 2E) and whole-brain (Fig. 2F) age gaps were regressed from all organ-specific age gaps and FGES was performed on the resulting residuals. The time interval between baseline and follow-up measurements was also regressed from baseline and follow-up age gaps, if appropriate (i.e., Fig. 2D). Influences inferred from FGES were represented using multiorgan networks, where each organ was denoted with a distinct network node. A directed edge was drawn from organ *X* to organ *Y* if and only if the age gap of *X* at baseline significantly influenced the follow-up age gap of *Y* (Fig. 2D) or rate of aging of *Y* (Fig. 2E,F), following false discovery rate (FDR) correction at 0.05 across the set of *J(J-1)/2* regression coefficients, where *J* denotes the total number of organ systems. If an edge was detected by FGES but the edge’s regression coefficient did not survive FDR correction, the edge was removed. Hence, drawing an edge required both statistical and causal evidence. In Fig. 2E,F, a positive regression coefficient provided evidence consistent with faster aging of organ *Y*, relative to organ *X* (i.e., a unit increase in the age of *X* predicted an increase of 𝛽 in the rate of aging of *Y*). In Fig. 2E,F, the baseline age gap node and rate of aging node were merged into a single node for each organ to provide a succinct network representation for visualization purposes. The Tetrad software package v6.8.1 (https://github.com/cmu-phil/tetrad) was used to perform the FGES heuristic, bootstrapping and parameter estimation. Default parameter settings for FGES were used (i.e., Chickering rule; BIC penalty discount: 0.5; T-depth: −1).

### Genetic, environmental and lifestyle factors

Telomere length (TL) shortening and human aging are linked^94^, and thus we tested for associations between organ-specific age gaps and the relative leucocyte TL (adjusted for technical parameters^95^) measured at baseline assessment, as well as genetic variants known to index leucocyte TL. Following an established method^96^, a polygenic score for leucocyte TL was computed for each individual based on nine single-nucleotide polymorphisms (SNPs) associated with leucocyte TL^97–99^. Larger polygenic scores associate with longer leucocyte TL, and vice versa. Age gaps for body and brain systems were also tested for associations with numerous environmental and lifestyle factors. We selected 158 variables that tapped individual differences in early life experience (e.g., birth weight, breastfed, adoption, maternal smoking and traumatic events), socio-demographics (e.g., education, neighborhood measure of deprivation, job status and parenting), lifestyle (e.g., smoking, alcohol intake, diet, exercise, sleep and e-device use), psychosocial (e.g., social support and mood status), local environmental exposures (e.g., air and noise pollution, greenspace and coastal proximity), general health (e.g., menstrual cycle, menopause, long-standing illness and disability, hearing, vision and falls) and cognitive ability. Individual variation in cognitive ability was measured using the cognitive age gap, inferred from the above-described cognitive age prediction model. Dummy variables were generated for categorical responses. Several variables were curated to enable more intuitive interpretation than the native UK Biobank coding. The curation procedure includes:

- Reponses indicating less than one unit in time, distance, frequency and quantity were originally coded as −10 and were recoded to 0 for all relevant variables, including food intake frequencies, time spent on watching television, using computer and driving, distance between home and job workplace etc.
- Individuals who did not provide a valid answer, originally coded as −3 (Prefer not to answer) or −1 (Do not know) were labelled as missing responses.
- An average weekly alcohol consumption (in UK standard units) was computed by combining information on each person’s response to questionnaire on weekly and monthly intake of a variety of beverage type, including red wine, white wine/champagne, beer/cider, spirits and fortified wine, consistent with previous literature^100, 101^. Specifically, weekly alcohol intake data was collected from individuals who indicated that they drink more often than once or twice a week, whereas monthly alcohol intake was collected from individuals who drink alcohol one to three times a month or on special occasions. The alcohol consumption of individuals who indicated that they never drink was set to zeros.
- Reponses to current tobacco smoking: 1-Yes, on most or all days; 2-Only occasionally; 0-No, were recoded to: 2-Yes, on most or all days; 1-Only occasionally; 0-No, so that higher scores denoted higher frequency of current tobacco smoking.
- Responses to past tobacco smoking were originally coded as: 1-Smoked on most or all days; 2-Smoked occasionally; 3-Just tried once or twice; 4-I have never smoked. Responses were thus reversed so that higher scores denoted higher frequency of past tobacco smoking.
- Individuals who smoked tobacco on most or all days in the past or current were labelled as “daily smokers”, notwithstanding varied definitions of smokers^102^.
- Time since stopped smoking was computed for past tobacco smokers by subtracting “age stopped smoking” from their chronological age at the assessment.
- Age started smoking in either past or current smokers was derived.
- An overall fruit and vegetable consumption (per day) was computed by summing fresh fruit, dried fruit, salad, cooked and raw vegetables intake^103^.
- Individuals who slept between 7 and 8 hours per night were labelled as had “good sleep duration”^104^.
- Reponses to facial aging were recoded to 1-younger than you are; 2-about your age; 3-older than you are, such that higher scores indicated older appearing face.
- Women who had no regular length of menstrual cycle were labelled as had irregular menstrual cycle.
- Women who were not sure if they have had menopause because of hysterectomy or other reasons were labelled as missing responses.
- Hearing, as measured by the speech reception threshold, was averaged for left and right ears.
- Visual acuity, as measured by the logarithm of the minimum angle of resolution (logMAR), was averaged for left and right eyes.

See table S6 for a full list of selected variables. The original UK Biobank Field IDs of variables were provided where applicable.

Partial correlation was used to test for associations between organ-specific age gaps and genetic, environmental and lifestyle factors, adjusting for chronological age and sex. Due to the very large sample sizes, statistical significance was Bonferroni corrected for genetic and environmental/lifestyle factors separately at p<0.004 (12 organ systems) and p<2.6×10^-5^ (158 factors × 12 organ systems = 1,896 tests), respectively. A minimum effect size threshold of |r|>0.05 was enforced to suppress weak associations for visualization purpose (Fig. 3). Of note, the Bonferroni correction was used to control the family-wise error when sample sizes were very large (>10,000). The false discovery rate (FDR) was controlled at 5% using the Benjamini-Hochberg procedure elsewhere.

### Health outcomes and clinical characterization

Diagnoses and medical conditions of participants were obtained through self-report (verbal interview at assessment centers, UK Biobank Field IDs: 20001; 20002) and health care records (e.g., hospital inpatient and primary care) from the UK National Health Services (NHS). Hospital inpatient records were summarized by distinct ICD (International Classification of Diseases and Related Health Problems)-9 and/or ICD-10 coded primary and/or secondary diagnoses for participants whose health outcomes resulted in a hospital admission. Summary inpatient diagnoses (Field IDs: 41270; 41271) in the July 2020 release were used in this study. Primary care data (Field ID: 42040) were sourced at record-level on 26 November 2020. Of note, primary care data in relation to clinical events were recorded by health professionals working at general practices using Read Codes Version 2 (Read V2) and Read Codes Clinical Terms Version 3 (Read CTV3). Diagnoses coded in Read were mapped to corresponding ICD codes according to the lookup table (*‘all_lkps_maps_v2.xlsx’*) provided by the UK Biobank (https://biobank.ndph.ox.ac.uk/showcase/showcase/auxdata/primarycare_codings.zip). For Read V2, the mapping was only performed when the Read code matched to a single ICD-9 or ICD-10 code. For Read CTV3, the mapping was only performed for Read code flagged as exact one-to-one mapping (‘E’) or target concept more general (‘G’) that had been completely refined (‘C’). Whereas self-report provided past and current medical conditions, health care records enabled a lifetime assessment of a participant’s health outcomes.

Based on self-report and health care records, we defined a healthy aging group and 16 clinical groups comprising individuals with a lifetime diagnosis of Parkinsonism, multiple sclerosis, stroke, dementia, depression, bipolar disorder, schizophrenia, ischemic heart disease, hypertensive diseases, chronic obstructive pulmonary disease (COPD), chronic kidney disease (CKD), diabetes, cirrhosis, osteoarthritis, osteoporosis and cancer. Each disease category was defined broadly with all causes and subtypes included. For example, the COPD group included self-reported COPD, emphysema/chronic bronchitis and emphysema; health care recorded ICD-9 coded emphysema (code 492) and chronic airway obstruction not elsewhere classified (496) and ICD-10 coded emphysema (code J43), MacLeod syndrome (J430), panlobular emphysema (J431), centrilobular emphysema (J432), other emphysema (J438), emphysema unspecified (J439), other COPD (J44), COPD with acute lower respiratory infection (J440), COPD with acute exacerbation unspecified (J441), other specified COPD (J448) and COPD unspecified (J449). Table S7 lists diagnostic codes related to each of the 16 disease categories. For each individual, the recorded date of diagnosis was compared across self-report and health care sources for each disease category, to determine whether the illness onset/diagnosis preceded or occurred after the baseline assessment of body and brain function. However, the earliest state of disease onset for some individuals may have not been captured because the data from general practitioners only covered approximately 45% of the UK Biobank cohort; in contrast to the more than 87% coverage of hospital inpatient records. To enable comparisons, the healthy aging group included individuals with no self-reported and/or healthcare documented lifetime chronic medical conditions. Proportions of healthy individuals included in the body (28,589/143,423=0.199) and brain (7,909/36,901=0.214) analyses subsamples are slightly greater than the proportion in the full sample (91,808/502,504=0.183; body: 𝜒^2^=203.43, *p*<2.2×10^-16^, brain: 𝜒^2^=228.01, *p*<2.2×10^-16^). This suggests that individuals for whom organ age was estimated are not necessarily representative of the full cohort. Demographic details of individuals comprising each defined clinical group are provided in table S8.

Individuals diagnosed with more than one disease category throughout their lifetime were assigned to multiple disease groups. As shown in fig. S7A, most individuals included in either body or brain aging analyses (n=169,109; 90,918 males) were linked with a single diagnostic category (n=52,113, 54.1%) and the proportion of individuals comorbid with 2 to 8 conditions was 28.3% (n=27,215), 11.6% (n=11,171), 4.1% (n=3,991), 1.4% (n=1,308), 0.4% (n=394), 0.1% (n=103) and 0.026% (n=25), respectively. The largest number of comorbidities was 9, in 4 individuals (0.0042%). Figure S7B,C shows a comorbidity network, representing a population-level lifetime co-occurrence of the 16 disease categories. The extent of comorbidity was quantified by correlating (Pearson correlation) the presence of categorical diagnoses (1-Yes or 0-No) across individuals for each sex. Permutation testing (n=10,000) was used to estimate p-values, and significant correlations (p<4.2×10^-4^) were Bonferroni corrected for (16×15)/2=120 disease pairs. Results were broadly consistent between females (fig. S7B) and males (fig. S7C) and that the 16 disease categories were parsed into two large comorbid groups, corresponding to major brain and body disorders. Interestingly, brain disorders were also comorbid with diseases primarily implicated in body organ systems, with stronger effect sizes observed in males. For example, depression was significantly associated with osteoarthritis, COPD and hypertensive diseases, and dementia was associated with CKD and stroke.

### Mortality risk prediction

Mortality data released on 4 March 2021 were used in this study. Individual mortality status (date of death) was determined using data linkages to national death registries in the UK, including NHS Digital (England and Wales) and NHS Central Register (Scotland). Data linkage procedures and steps in data cleaning and validation are described in detail in the central UK Biobank death linkage documentation: (https://biobank.ndph.ox.ac.uk/showcase/showcase/docs/DeathLinkage.pdf).

Mortality was confirmed in 8,109 (age of death 42-83 years, mean 69.6±7.3, 5,670 males) and 330 (age of death 53-82 years, mean 70.7 ± 6.4, 203 males) individuals after baseline assessment of body and brain function, respectively. We first compared the mean age gap between deceased and non-deceased individuals for each organ system using the two-sample *t*-test. Cox proportional hazards regression was then used to estimate the risk of mortality associated with organ-specific age gaps. Lastly, we developed a logistic model using 10-fold cross-validation to predict an individual’s 5- and 10-year survival and premature mortality based on organ-specific age gaps. The risk of mortality associated with body and brain age were estimated separately because of the time difference in assessments (Fig. 2C).

The Cox proportional hazards model was applied under the assumption that mortality hazard ratios in relation to organ age gaps do not change over time for any individual. Therefore, the estimated hazard ratio represented the relative risk of death for each unit increase in age gap, compared to the baseline hazard, which was defined as the mean age gap across individuals. To enable comparisons, each organ age gap was first standardized by mean and standard deviation (SD). Two Cox regression models were then formulated, where one model estimated the mortality hazard ratios per one SD increase in organ-specific age gap, adjusting for sex and chronological age (standardized), and the second model further adjusted for existing diagnoses, general health (i.e., long-standing illness) and key lifestyle factors including smoking, exercise, socioeconomic inequality (deprivation) and tertiary education. Key lifestyle factors were selected based on i) significant associations with body/brain age gap (ranked within the top 20 out of 158 measures); and ii) no missing responses among deceased individuals. Survival was ascertained up to 13.41 (mean 7.47±3.2) and 6.07 (mean 2.47±1.49) years after baseline assessment of body and brain function, respectively. Non-deceased individuals were right-censored, where survival duration was calculated as days between the date of body or brain assessment and the date of mortality ascertainment (4 March 2021).

Finally, using 10-fold cross-validation, logistic models were trained to predict the probability of an individual’s survival time and premature mortality based on body age gaps. A predictive model was not developed for brain age gaps, given the lack of evidence for a link between advanced brain age and mortality risk, as estimated from Cox regression (fig. S11B,C). For survival time (T) prediction, a nominal logistic regression was fitted to classify whether an individual was deceased within 5 years (T<5) or had lived more than 5 years (T≥5, deceased or non-deceased) after assessment of body function. The same model was fitted for 10-year survival time. All non-deceased individuals had lived more than 10 years (minimal survival time: 10.43 years) after body function assessment. Similarly, logistic models were developed to predict an individual’s premature death. Premature death is typically defined as death occurring before the mean age of death in a certain population, which is approximately 75 years in the UK^105^. Death before age 70 years was also considered based on the mean age of death of the current UK Biobank cohort (n=34,003, mean age of death 69.6±7.4 years). Prediction models were estimated based on five groups of predictors to assess the extent to which body age gaps improved prediction of survival and premature death beyond established predictors, including chronological age, sex, existing disease diagnoses and key lifestyle factors. The six models were as below:

- Model 1: chronological age and sex.
- Model 2: chronological age, sex and eight body system ages.
- Model 3: chronological age, sex and existing diagnoses of the 16 disease categories.
- Model 4: chronological age, sex, eight body age gaps and existing diagnoses of the 16 disease categories.
- Model 5: chronological age, sex, eight body age gaps, existing diagnoses of the 16 disease categories, general health and key lifestyle factors included in the Cox regression analysis.
- Model 6: chronological age, sex, existing diagnoses of the 16 disease categories, general health and key lifestyle factors included in the Cox regression analysis.

Chronological age and body age gaps were standardized by mean and standard deviation before model training. The area under the curve (AUC) of the receiver operating characteristic curve was used to quantify prediction accuracy. Confidence intervals were estimated with bootstrapping (100 samples).

## Data Availability

Researchers can register to access all data used in this study via the UK Biobank Access Management System (https://bbams.ndph.ox.ac.uk/ams/).

## Acknowledgments

This research has been conducted using data from UK Biobank (https://www.ukbiobank.ac.uk/), a major biomedical database. We are grateful to UK Biobank for making the data available, and to all study participants, who generously donated their time to make this resource possible. Some of the data used in the preparation of this article were obtained from the Australian Imaging Biomarkers and Lifestyle flagship study of ageing (AIBL), funded by the Commonwealth Scientific and Industrial Research Organisation (CSIRO) which was made available at the ADNI database (https://adni.loni.usc.edu/). The AIBL researchers contributed data but did not participate in analysis or writing of this report. AIBL researchers are listed at: https://aibl.csiro.au/. Some of the data used in preparation of this article were obtained from the Alzheimer’s Disease Neuroimaging Initiative (ADNI) database (https://adni.loni.usc.edu/). As such, the investigators within the ADNI contributed to the design and implementation of ADNI and/or provided data but did not participate in analysis or writing of this report. A complete listing of ADNI investigators can be found at: http://adni.loni.usc.edu/wp-content/uploads/how_to_apply/ADNI_Acknowledgement_List.pdf. Some of the data collection and sharing for this project was funded by the Alzheimer’s Disease Neuroimaging Initiative (ADNI) (National Institutes of Health Grant U01 AG024904) and DOD ADNI (Department of Defense award number W81XWH-12-2-0012). ADNI is funded by the National Institute on Aging, the National Institute of Biomedical Imaging and Bioengineering, and through generous contributions from the following: AbbVie, Alzheimer’s Association; Alzheimer’s Drug Discovery Foundation; Araclon Biotech; BioClinica, Inc.; Biogen; Bristol-Myers Squibb Company; CereSpir, Inc.; Cogstate; Eisai Inc.; Elan Pharmaceuticals, Inc.; Eli Lilly and Company; EuroImmun; F. Hoffmann-La Roche Ltd and its affiliated company Genentech, Inc.; Fujirebio; GE Healthcare; IXICO Ltd.;Janssen Alzheimer Immunotherapy Research & Development, LLC.; Johnson & Johnson Pharmaceutical Research & Development LLC.; Lumosity; Lundbeck; Merck & Co., Inc.;Meso Scale Diagnostics, LLC.; NeuroRx Research; Neurotrack Technologies; Novartis Pharmaceuticals Corporation; Pfizer Inc.; Piramal Imaging; Servier; Takeda Pharmaceutical Company; and Transition Therapeutics. The Canadian Institutes of Health Research is providing funds to support ADNI clinical sites in Canada. Private sector contributions are facilitated by the Foundation for the National Institutes of Health (https://www.fnih.org/). The grantee organization is the Northern California Institute for Research and Education, and the study is coordinated by the Alzheimer’s Therapeutic Research Institute at the University of Southern California. ADNI data are disseminated by the Laboratory for Neuro Imaging at the University of Southern California. We thank Stephen M. Smith (University of Oxford) and Didac Vidal-Piñeiro (University of Oslo) for their feedback and discussion on data analyses. Y.E.T was supported by the Mary Lugton Postdoctoral Fellowship. A.Z. was supported by the National Health and Medical Research Council (NHMRC) grant (APP1142801 and APP118153). V.C. was supported by the NHMRC grant (APP1177370). M.B. was supported by the NHMRC grant (IG2008612, APP1095227 and APP1092623).

## Author contributions

Conceptualization: Y.E.T, A.Z, V.C, M.B

Methodology: Y.E.T, A.Z, M.B

Investigation: Y.E.T, A.Z

Visualization: Y.E.T, A.Z

Project administration: Y.E.T

Supervision: A.Z, V.C

Writing – original draft: Y.E.T, A.Z

Writing – review & editing: Y.E.T, A.Z, V.C, M.B, A.B.M, N.T.L

## Data and materials availability

Matlab (Natick, Massachusetts: The MathWorks Inc.) code for conducting the core analyses is available on GitHub (https://github.com/yetianmed/BioAge). Structural equation modeling was performed using the Tetrad software package v6.8.1 (https://github.com/cmu-phil/tetrad). The organ images shown in Figure 1 were created with BioRender.com. Other figures were created using visualization routines in Matlab). Participant age gaps for all body and brain systems estimated in this study will be returned to the UK Biobank to strengthen the resource and facilitate access to other researchers for future research. Researchers can register to access all data used in this study via the UK Biobank Access Management System (https://bbams.ndph.ox.ac.uk/ams/).

## Supplementary Materials

**Fig. S1.**
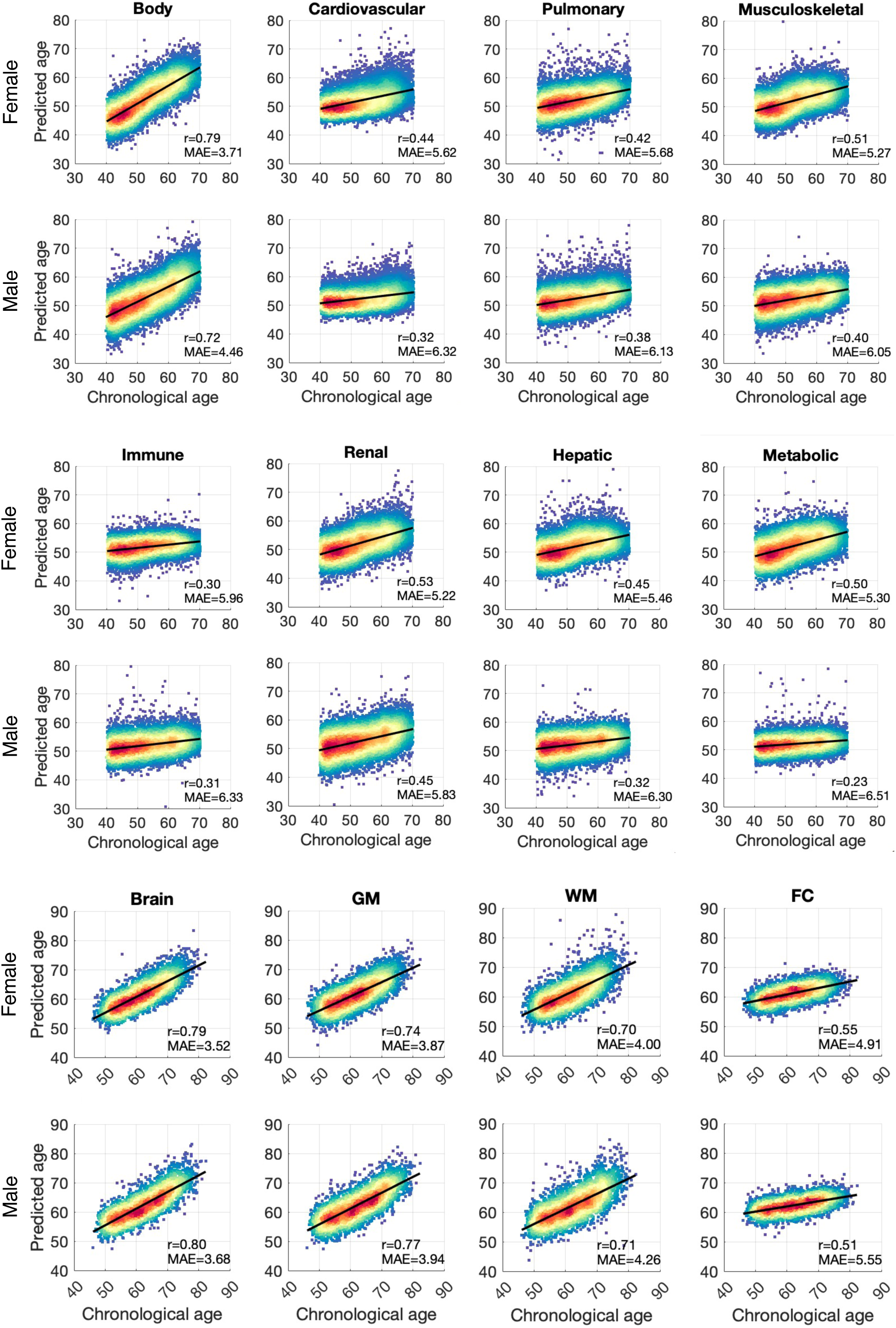
Age prediction accuracy. Scatter plots show associations between chronological and predicted age for prediction models based on body and brain phenotypes as well as phenotypes pertaining to each individual organ system. Lines of best fit indicated with solid black lines. r: Pearson correlation coefficients; MAE: mean absolute error.

**Fig. S2.**
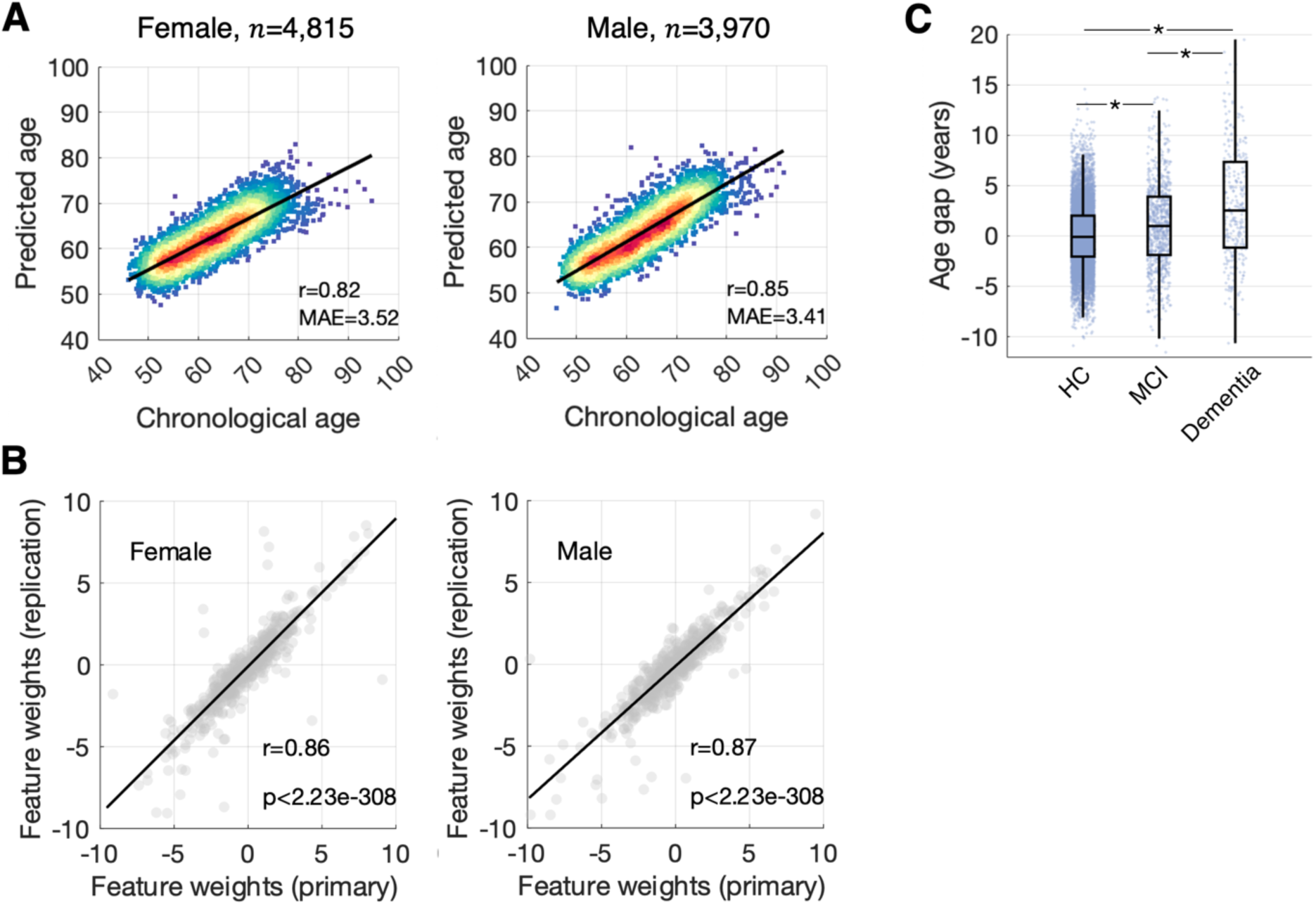
Replication of predictive models for brain gray matter age. **(A)** Scatter plots show associations between chronological age and predicted age for prediction model based on brain gray matter phenotypes in a combined group of healthy individuals from the Australian Imaging, Biomarkers and Lifestyle Flagship Study of Ageing (AIBL, n=396, 154 males), the Alzheimer’s Disease NeuroImaging Initiative (ADNI, n=467, 192 males) and the UK Biobank (n=7,922, 3,624 males). Lines of best fit indicated with solid black lines. n: training sample size; r: Pearson correlation coefficients; MAE: mean absolute error. **(B)** Scatter plots show associations between gray matter feature weights estimated from the original age prediction model (primary) and the re-trained model using the replication cohort. Lines of best fit indicated with solid black lines. r: Pearson correlation coefficients. **(C)** Gray matter age (i.e., age gap) in individuals diagnosed with mild cognitive impairment (MCI, n=780, mean age gap=1.07±4.25 years) and dementia (n=284 mean age gap=3.19±6.13 years), compared to healthy individuals (HC). The mean age gap significantly differs across the three groups (*F*-statistic=157.49, *p*=4.71×10^-68^). Asterisks indicate significant between-group differences, adjusting for chronological age and sex (MCI vs HC, *t*=10.39, *p*=3.56×10^-25^; dementia vs HC, *t*=16.94, *p*<2.23×10^-308^; MCI vs dementia: *t*=10.76, *p*=1.11×10^-25^).

**Fig. S3.**
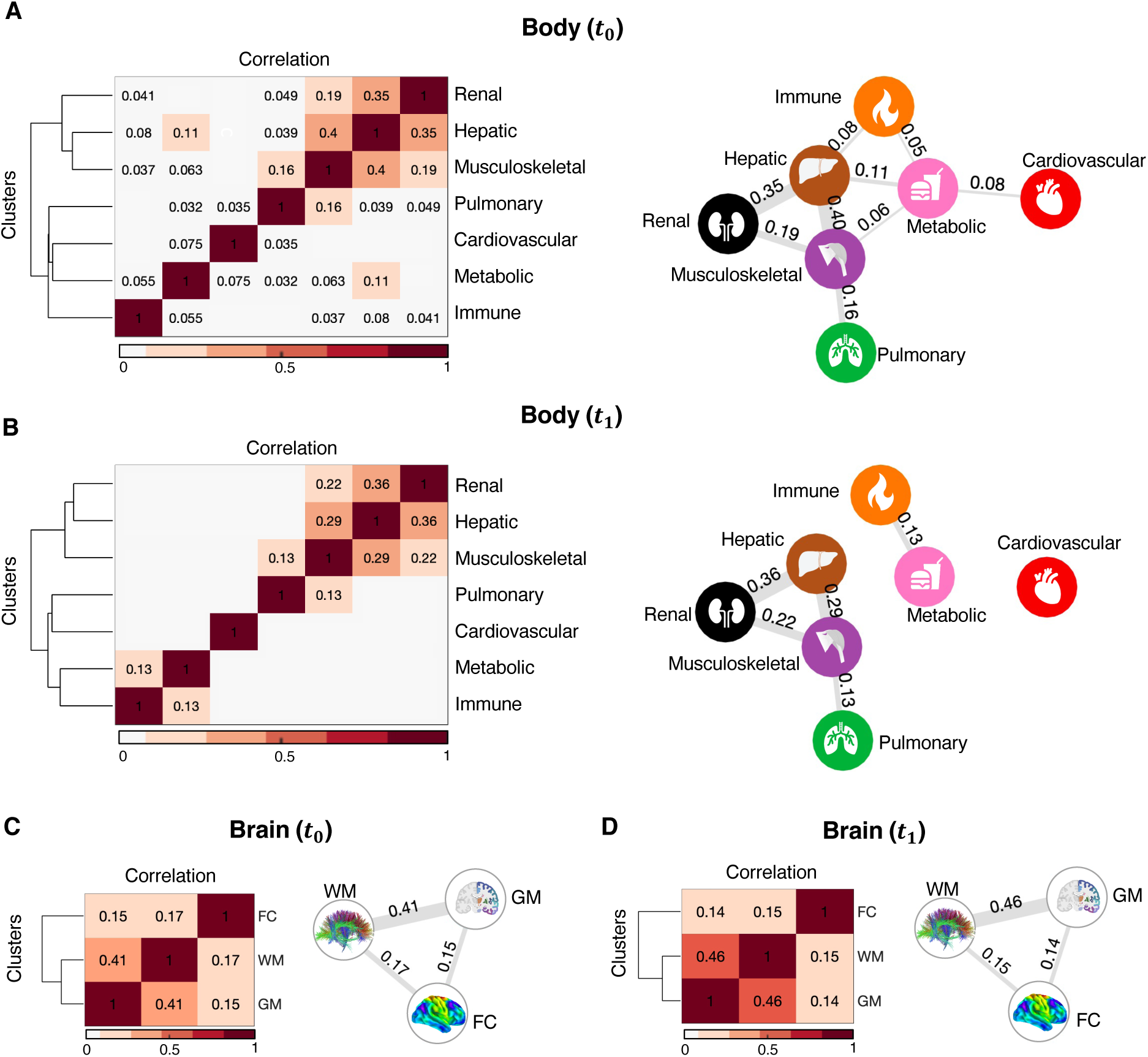
Synchrony among organ-specific age gaps. **(A)** Synchrony in biological ages between each pair of body systems at baseline assessment was estimated using partial correlation, adjusting for sex and chronological age. Correlation coefficients of significant pairs of correlations (p<0.002, Bonferroni corrected for 21 pairs) are indicated in the matrix (left) and also visualized as a graph (right). In the graph, each node represents one of the 7 body organs and the edges between them indicate correlations. Edge thicknesses are proportional to correlation coefficients. Edges are suppressed for small effect sizes (|r|<0.05) **(B)** Same as (A) but shows the correlations at follow-up assessment. Body systems can be differentiated into two groups based on interorgan synchrony in age gaps (Group I: renal, hepatic, musculoskeletal; Group II: pulmonary, cardiovascular, metabolic, immune). **(C)** & **(D)** Same as (A) & (B) but the synchrony in age gaps is shown for different brain systems at baseline and follow-up assessment respectively. Biological age is most strongly synchronized between white and gray matter, whereas functional connectivity is only weakly synchronized with other brain systems (Bonferroni corrected for 3 correlations, p<0.017). GM, gray matter; WM, white matter; FC, functional connectivity. Ward’s linkage clustering was used to determine the reordering and the cluster tree shown.

**Fig. S4.**
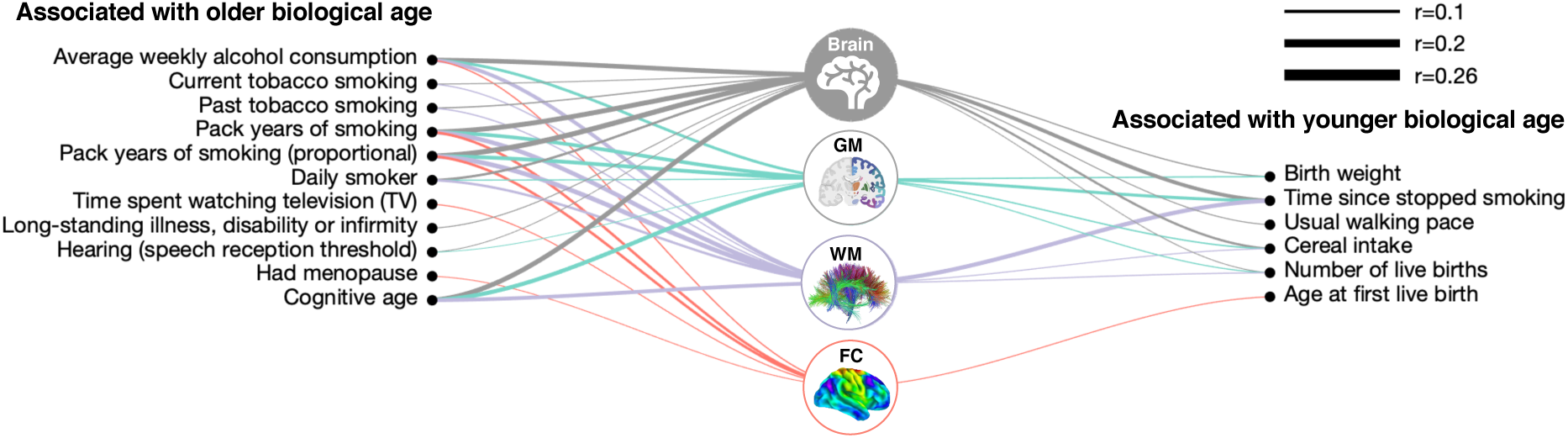
Associations between environmental/lifestyle factors and brain age gaps. Icons represent specific brain systems for which biological age was estimated. Links are shown between environmental/lifestyle factors significantly associated with age gaps of specific brain systems (p<2.6×10^-5^, Bonferroni corrected for 158 factors × 12 body and brain systems = 1896 tests). Links are suppressed for small effect sizes (|r|<0.05). Left (right) list comprises factors associated with brain systems that appear older (younger) than same-aged peers. Partial correlation was used to test for associations between brain age gaps and environmental/lifestyle factors, adjusting for chronological age and sex. Link widths are proportional to correlation coefficients. Results for other organ systems are shown in Fig. 3 in the main text. GM, gray matter; WM, white matter; FC, functional connectivity.

**Fig. S5.**
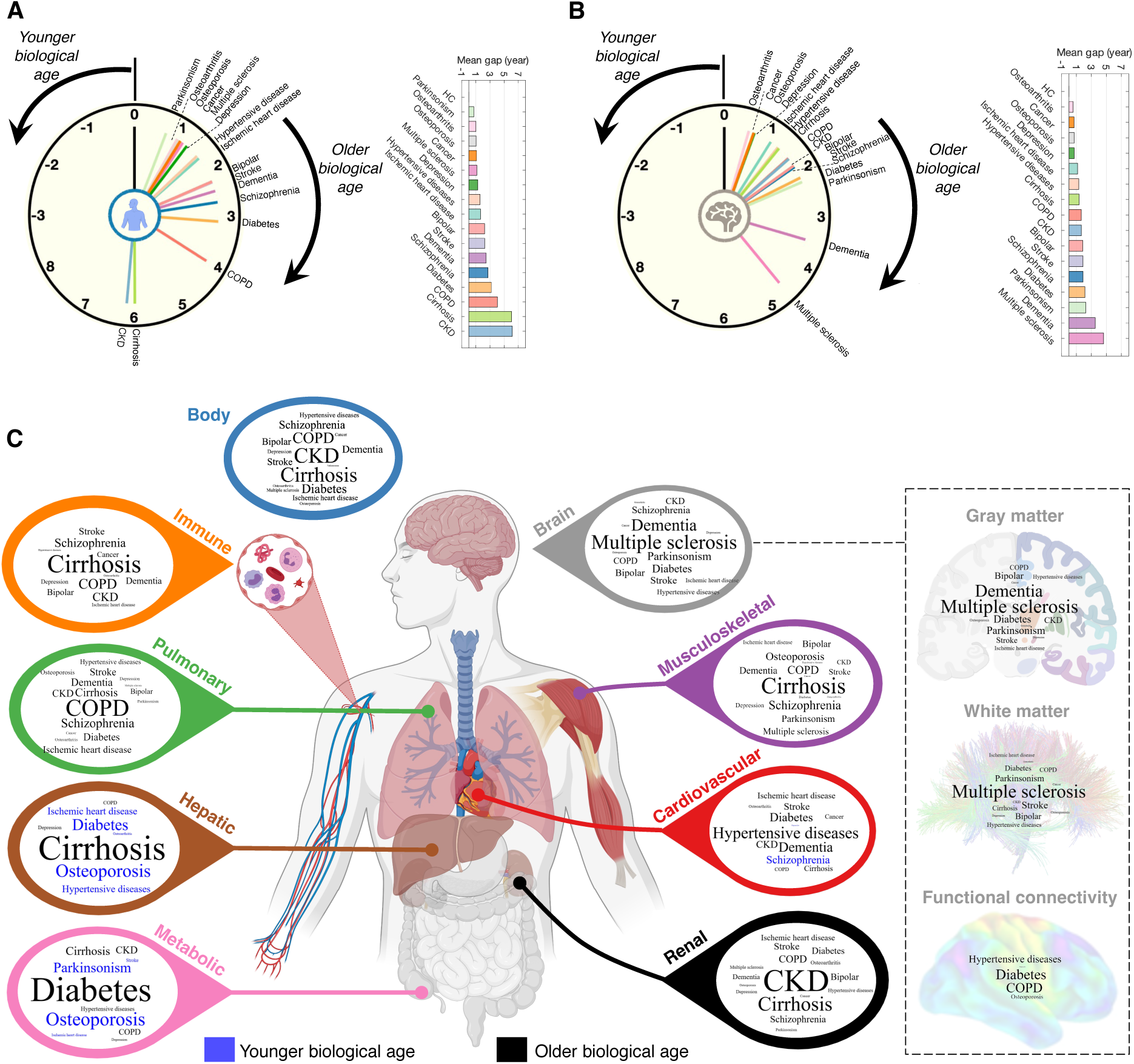
Relationship between chronic disease and organ-specific biological age. **(A)** A clock face represents the extent of body aging for 16 disease categories. Body age is older (younger) in a clockwise (anticlockwise) direction, with a body age gap of zero at the 12 o’clock position. Bar plot shows the mean body age gap in each disease, sorted from the smallest to the largest value. **(B)** Same as panel (A) but shows the mean brain age gap across disease. Dashed arm indicates replication dementia cohort. **(C)** Word-cloud representation. The font size was normalized according to the mean age gap across the 16 disease groups within each organ system. Diseases for which organs appear older than chronological age (gap>0) are colored black, whereas diseases for which organs appear younger (gap<0) are colored blue. COPD, chronic obstructive pulmonary disease; CKD, chronic kidney disease. Organ image was created with BioRender.com.

**Fig. S6.**
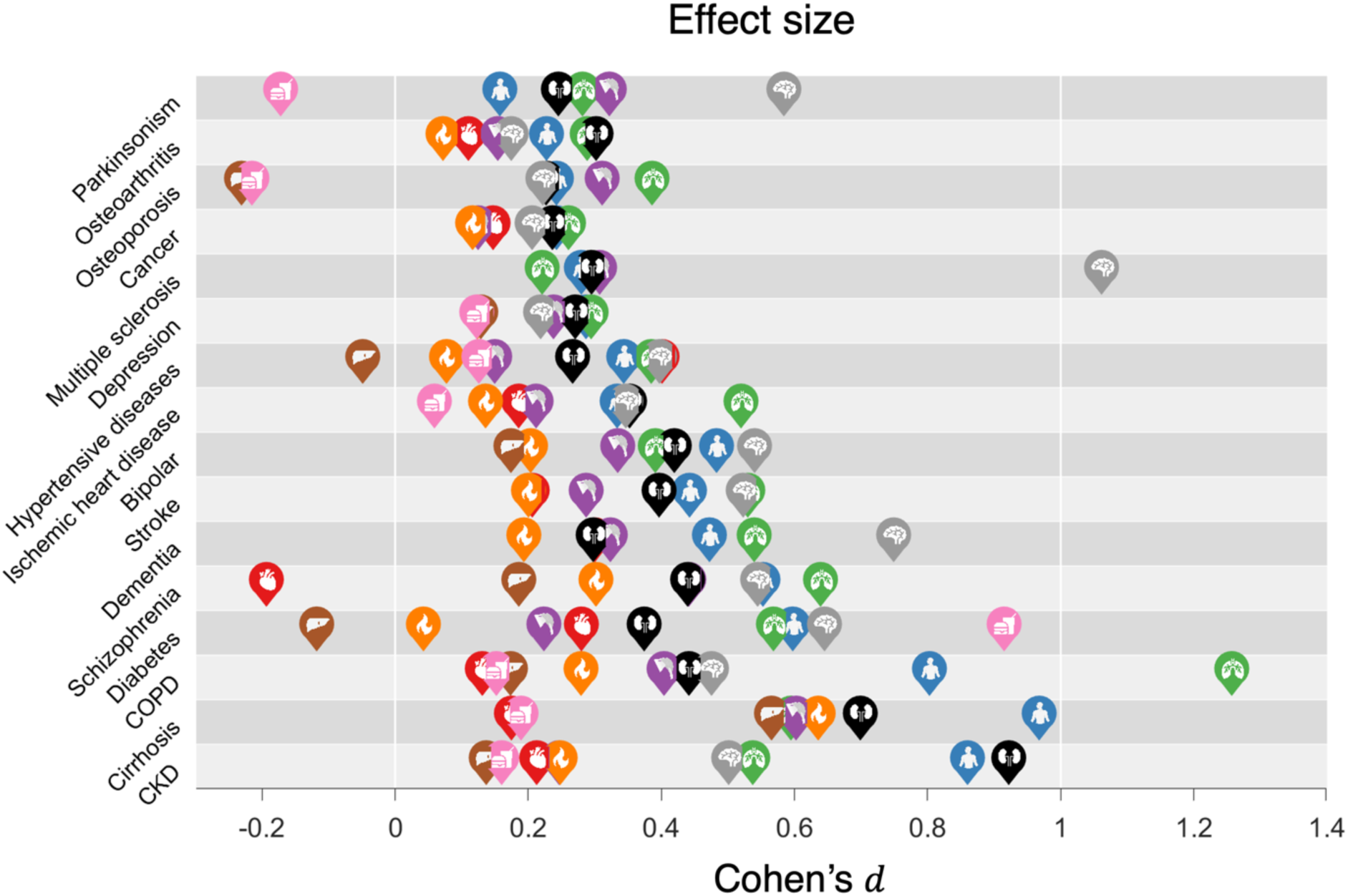
Effect size of body and brain age in chronic disease. Effect sizes of differences in organ-specific age gaps between each disease category and healthy comparison group were quantified using the Cohen’s *d*. The Cohen’s d value was multiplied by the sign of the mean between-group difference in age gap. Icons representing body systems and organs are positioned to indicate the effect size for each disease category. Icons are not shown for organs with mean age gaps that do not significantly differ from zero (p<2.6 × 10^-4^, Bonferroni corrected, Fig. 4A). Disease categories are ordered from top to bottom according to increased mean body age gaps as shown in Fig. 4B.

**Fig. S7.**
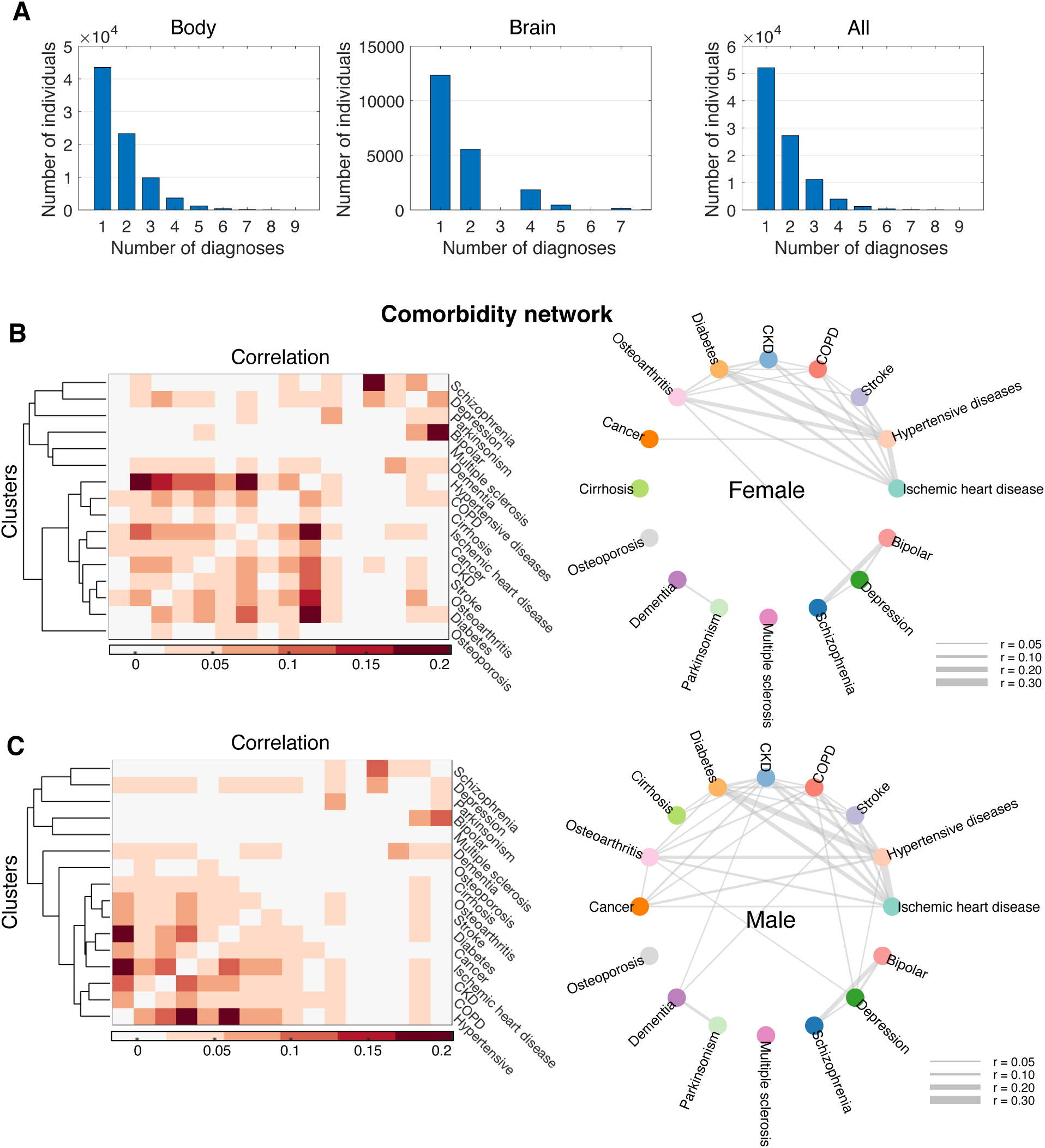
Disease comorbidity. (**A**) Bar plots show the number of lifetime comorbid diagnoses for individuals who completed assessment of body (left) or, brain (middle) function and all individuals (right). (**B**) Comorbidity network for females. The Pearson correlation coefficient was used to quantify the extent of lifetime comorbidity between each pair of disease categories. Permutation testing (n=10,000) was used to estimate p-values and significant correlations were Bonferroni corrected for (16×15)/2=120 disease pairs (p<4.2×10^-4^). Non-significant correlations were suppressed from the correlation matrix (left) and the network graph (right). Edge thickness is modulated by correlation coefficients. (**C**) Same as (D) but for males. Also see Methods.

**Fig. S8.**
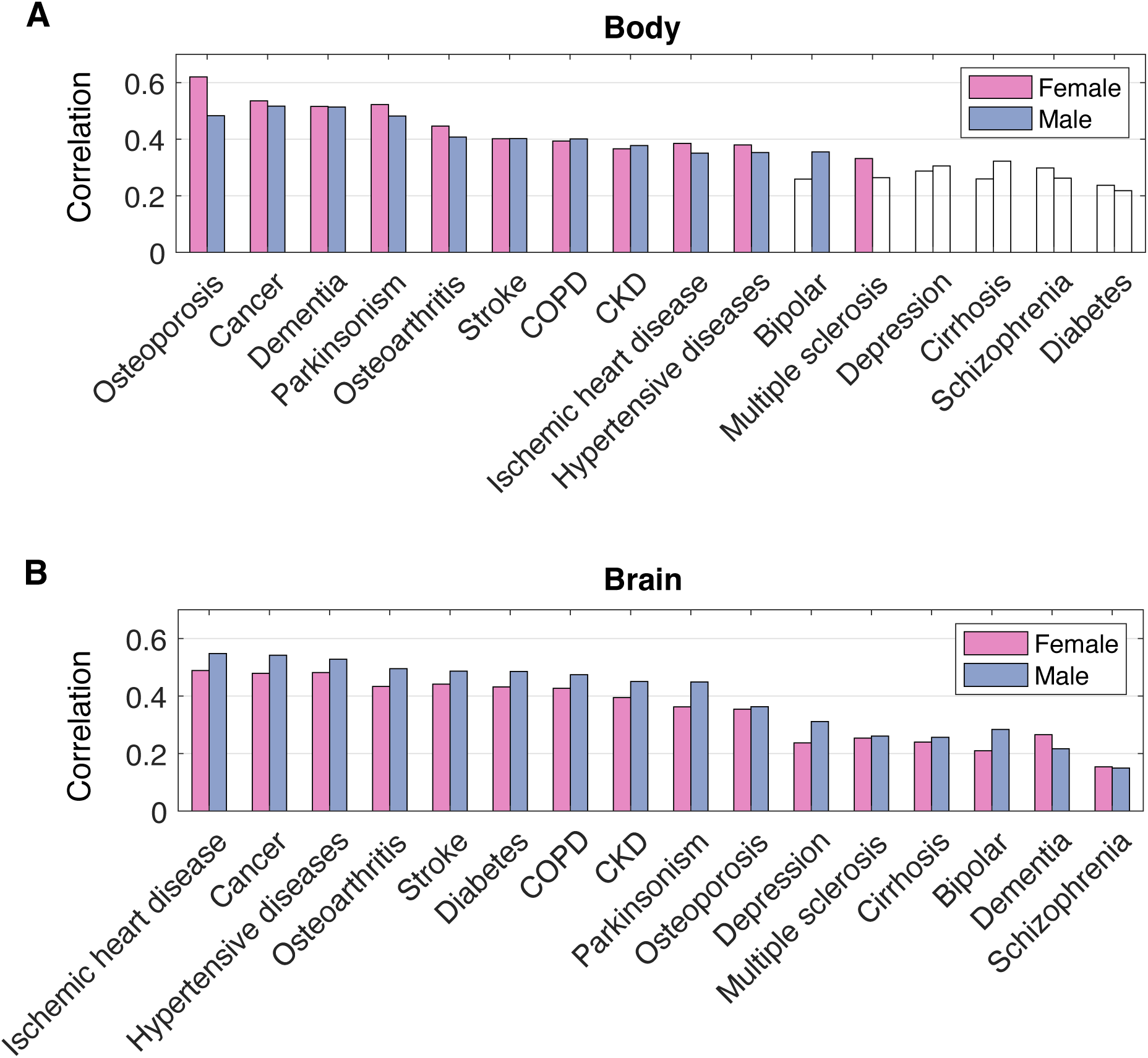
Associations between aging and disease-related changes in body and brain health. (**A**) Bar plots show Pearson correlation coefficients between regression coefficients (i.e., feature weights) of the age prediction model and between-group differences (health vs disease) in the body features (n=79), stratified by each disease category (p<0.003, Bonferroni corrected for 16 disease groups). Bar plots are colored white for non-significant correlations. (**B**) Correlations between the regression coefficients in age prediction and the between-group differences (health vs disease) in the brain features (n=2,309), stratified by each disease category.

**Fig. S9.**
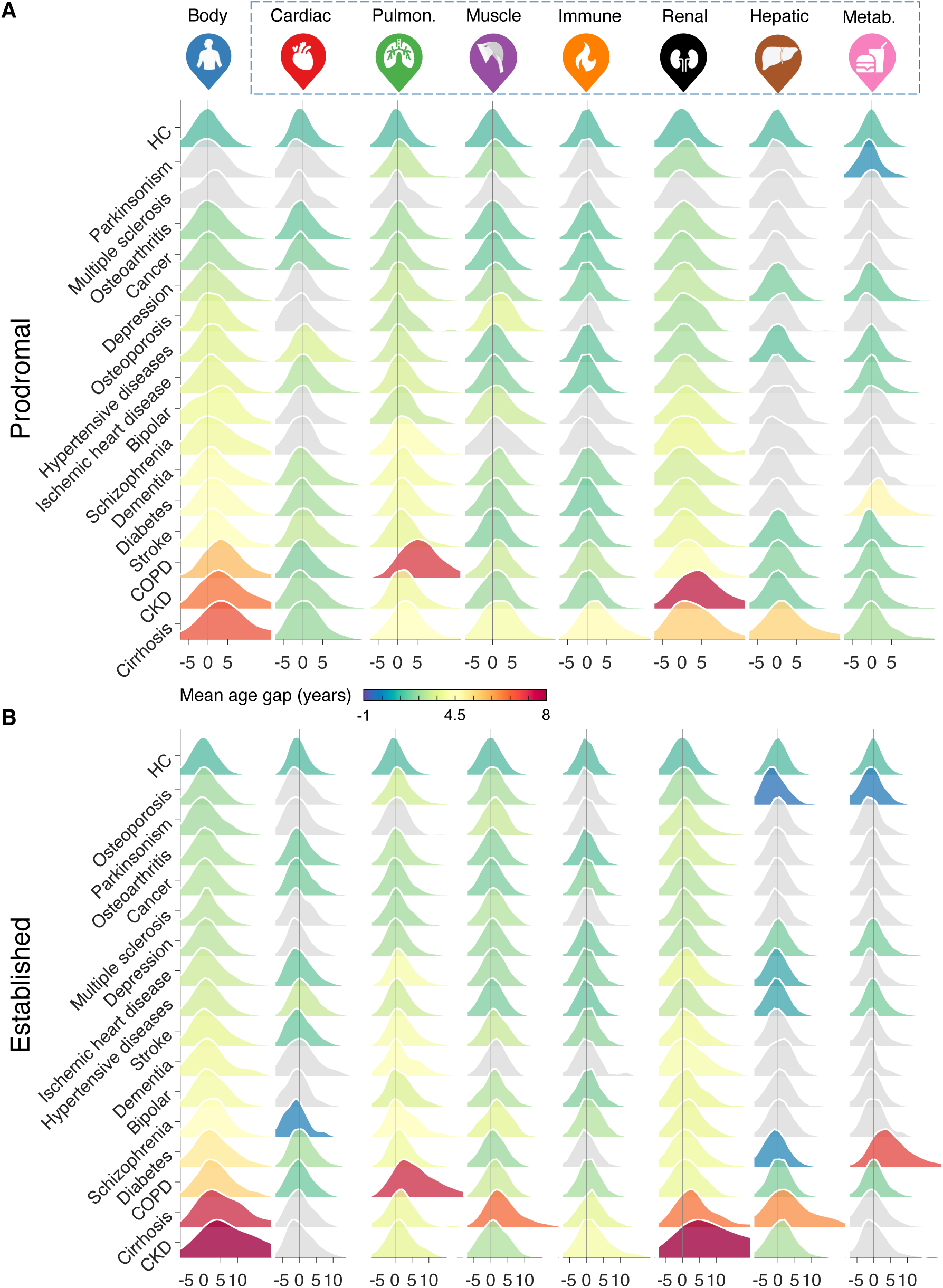
Biological organ age in chronic disease at different illness stages. (**A**) Distribution of body age gaps (columns) for 16 disease categories (rows) in individuals at prodromal stage, compared to healthy individuals (HC, first row). Distributions are colored according to disease- and organ-specific mean age gaps. Colored distributions have a mean that significantly differs from the healthy group (p<3.9×10^-4^, Bonferroni corrected for 16 disease categories × 8 body systems = 128 tests). Distributions colored gray have a mean that is not significantly different from the healthy group. (**B**) Same as (A) but in individuals with established diagnosis. Prodromal groups for brain imaging data were insufficient to investigate the impact of disease progression on brain age.

**Fig. S10.**
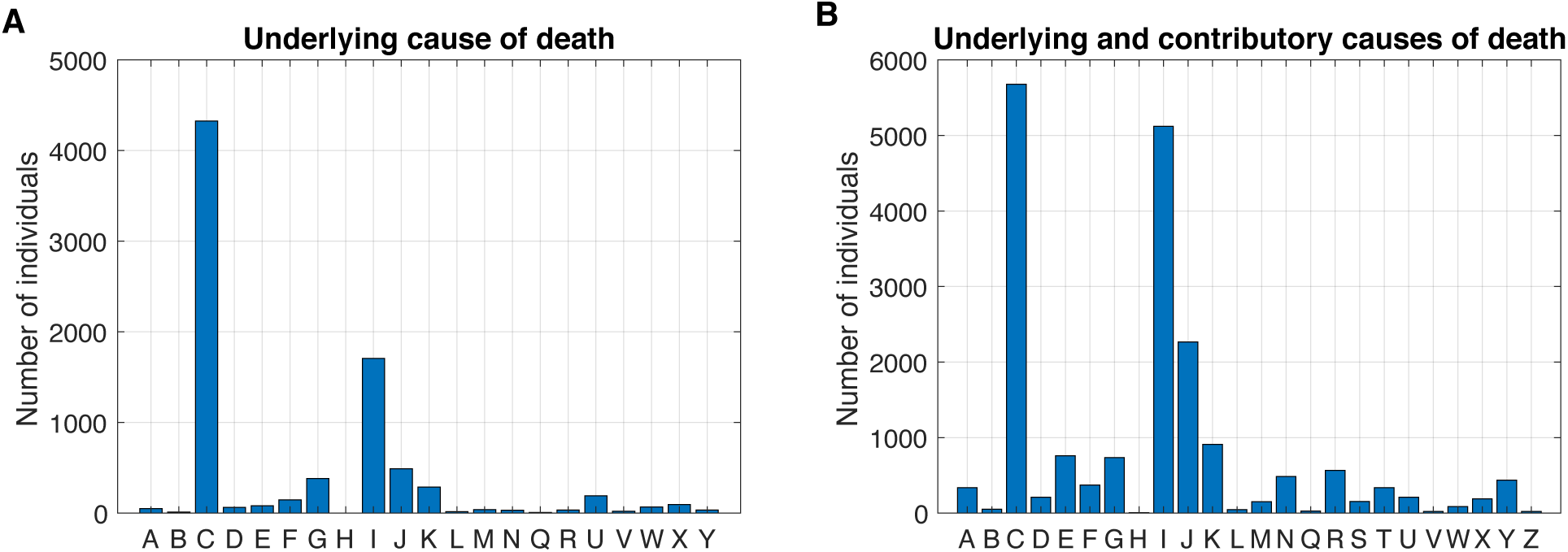
Causes of death. Underlying (**A**) and/or contributory (**B**) causes of death among 8,109 deceased individuals. Each capital letter (x-axis) represents a broad cause of death category coded by the International Statistical Classification of Diseases and Related Health Problems 10th Revision (ICD-10). A-B, certain infectious and parasitic diseases; C, malignant neoplasms (cancer); D, benign and other neoplasms, diseases of the blood and blood-forming organs and certain disorders involving the immune mechanism; E, endocrine, nutritional and metabolic diseases; F, mental and behavioral disorders; G, diseases of the nervous system; H, diseases of the eye and adnexa, ear and mastoid process; I, diseases of the circulatory system; J, diseases of the respiratory system; K, disease of the digestive system; L, diseases of the skin and subcutaneous tissue; M, diseases of the musculoskeletal system and connective tissue; N, diseases of the genitourinary system; Q, congenital malformations, deformations and chromosomal abnormalities; R, systems, signs and abnormal clinical and laboratory findings, not elsewhere classified; S-T, injury, poisoning and certain other consequences of external causes; U, provisional assignment of new diseases of uncertain etiology or emergency use; V-Z, external causes of morbidity and mortality.

**Fig. S11.**
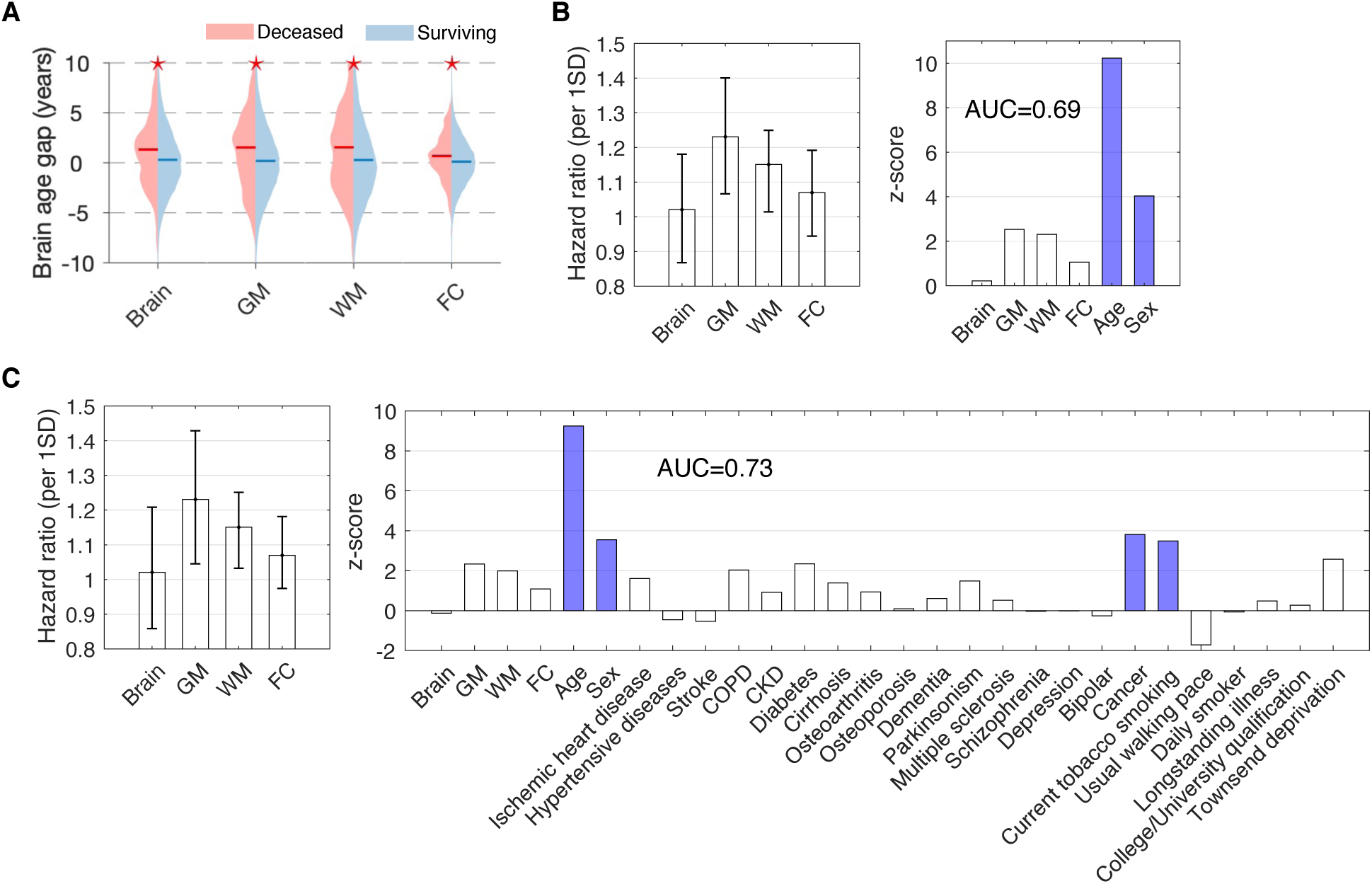
Brain age and the risk of mortality. (**A**) Brain age gaps in deceased (n=330) compared to non-deceased (n=36,571) individuals on 4 March 2021. Asterisks indicate significant between-group differences (two-sample t-test, p<0.0125, Bonferroni corrected for 4 brain systems). (**B**) The risk of mortality associated with brain age, after controlling for chronological age and sex. This was estimated using Cox proportional hazard regression, where survival durations were right censored for surviving individuals. Bar plots show the hazard ratio per one standard deviation (SD) change in the age gap (left) and z-scores for each brain system in the regression (right). Confidence intervals (95%) estimated with bootstrapping (n=100). Colored bars indicate risk factors with significant hazard ratios (Bonferroni correction at p<0.05/6=0.008). Gray (p=0.01, uncorrected) and white (p=0.02, uncorrected) matter age showed nominally significant hazard ratios. AUC: area under curve. (**C**) Same as (B) but existing disease diagnoses, general health and key lifestyle factors included in the regression. Gray (p=0.02) and white (p=0.04) matter age was nominally significantly associated with mortality risk after controlling for these factors but did not survive Bonferroni correction at p<0.05/29=0.0017. GM, gray matter; WM, white matter; FC, functional connectivity.

**Fig. S12.**
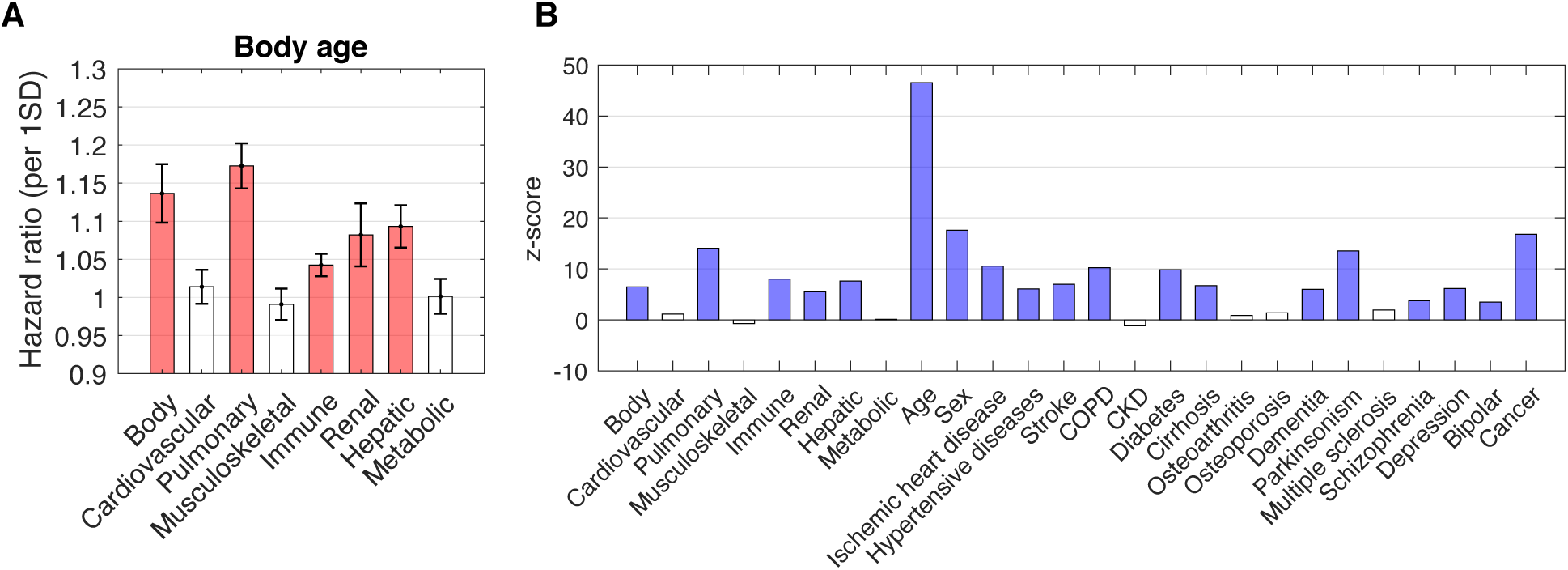
Body age and the risk of mortality. (**A**) Bar plots show mortality hazard ratios per one standard deviation (SD) change in organ-specific age gap and (**B**) corresponding z-scores. Hazard ratios estimated using Cox proportional hazard regression, where survival durations were right censored for surviving individuals. Chronological age, sex and existing diagnoses included as confounds. Confidence intervals (95%) estimated with bootstrapping (n=100). Colored bars indicate body systems with significant hazard ratios (p<0.0019, Bonferroni corrected for 26 dependent variables). AUC: area under curve.

**Fig. S13.**
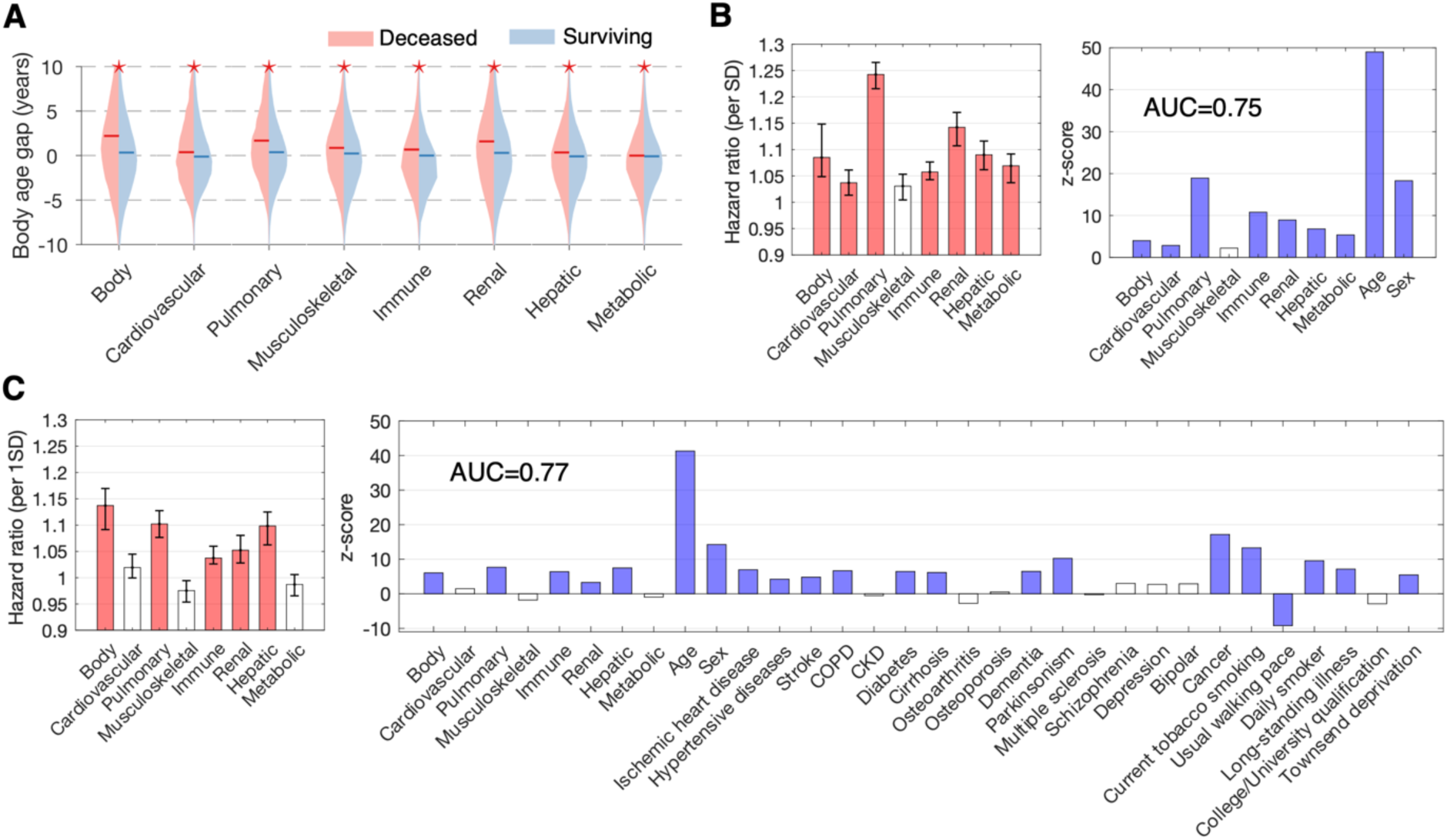
Body age and the risk of mortality. Supplementary analyses using mortality data ascertained before the first recorded death from coronavirus disease (COVID-19) in the UK on 6 March 2020^33^. Survival after baseline body function assessment was ascertained up to 12.32 years (n=6,986, age of death 42-82 years, mean 68.9±7.2, 4,899 males). **(A)** Body age gaps in deceased (n=6,986) compared to non-deceased (n=136,437) individuals. Asterisks indicate significant between-group differences, controlling for chronological age and sex (p<0.0056, Bonferroni corrected). **(B)** Bar plots show mortality hazard ratio per one standard deviation (SD) increase in organ-specific age (left) and corresponding z-scores (right). Chronological age and sex are included in the regression. Confidence intervals (95%) estimated with bootstrapping (n=100). Colored bars indicate organs with significant hazard ratios (p<0.005, Bonferroni corrected for 10 dependent variables). AUC: area under curve. **(C)** Same as panel (B), but existing disease diagnoses, general health and key lifestyle factors included in the regression. Colored bars indicate organs with significant hazard ratios (p<0.0015, Bonferroni corrected for 32 dependent variables).

**Fig. S14.**
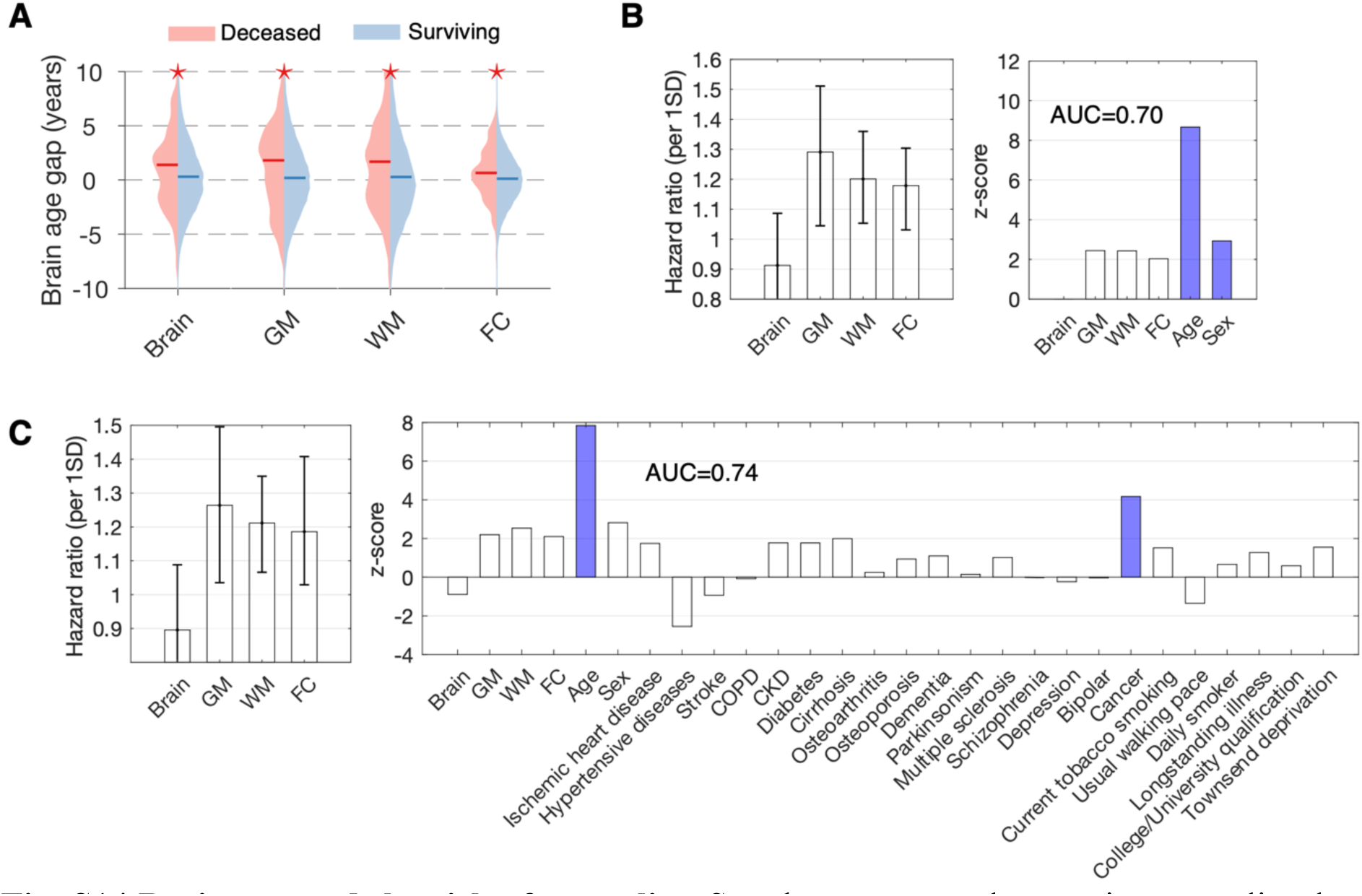
Brain age and the risk of mortality. Supplementary analyses using mortality data ascertained before the first recorded death from COVID-19 in the UK on 6 March 2020^33^. Survival after baseline brain function assessment was ascertained up to 5.22 years (n=203, age of death 53-81 years, mean 70.4±6.1, 124 males). **(A)** Brain age gaps in deceased (n=203) compared to non-deceased (n=36,689) individuals. Asterisks indicate significant between-group differences, controlling for chronological age and sex (p<0.0056, Bonferroni corrected). (**B**) Bar plots show mortality hazard ratio per one standard deviation (SD) increase in organ-specific age (left) and corresponding z-scores (right). Chronological age and sex are included in the regression. Confidence intervals (95%) estimated with bootstrapping (n=100). Colored bars indicate organs with significant hazard ratios (p<0.008, Bonferroni corrected for 6 dependent variables). AUC: area under curve. **(C)** Same as panel (B), but existing disease diagnoses, general health and key lifestyle factors included in the regression. Colored bars indicate organs with significant hazard ratios (p<0.0017, Bonferroni corrected for 29 dependent variables).

**Fig. S15.**
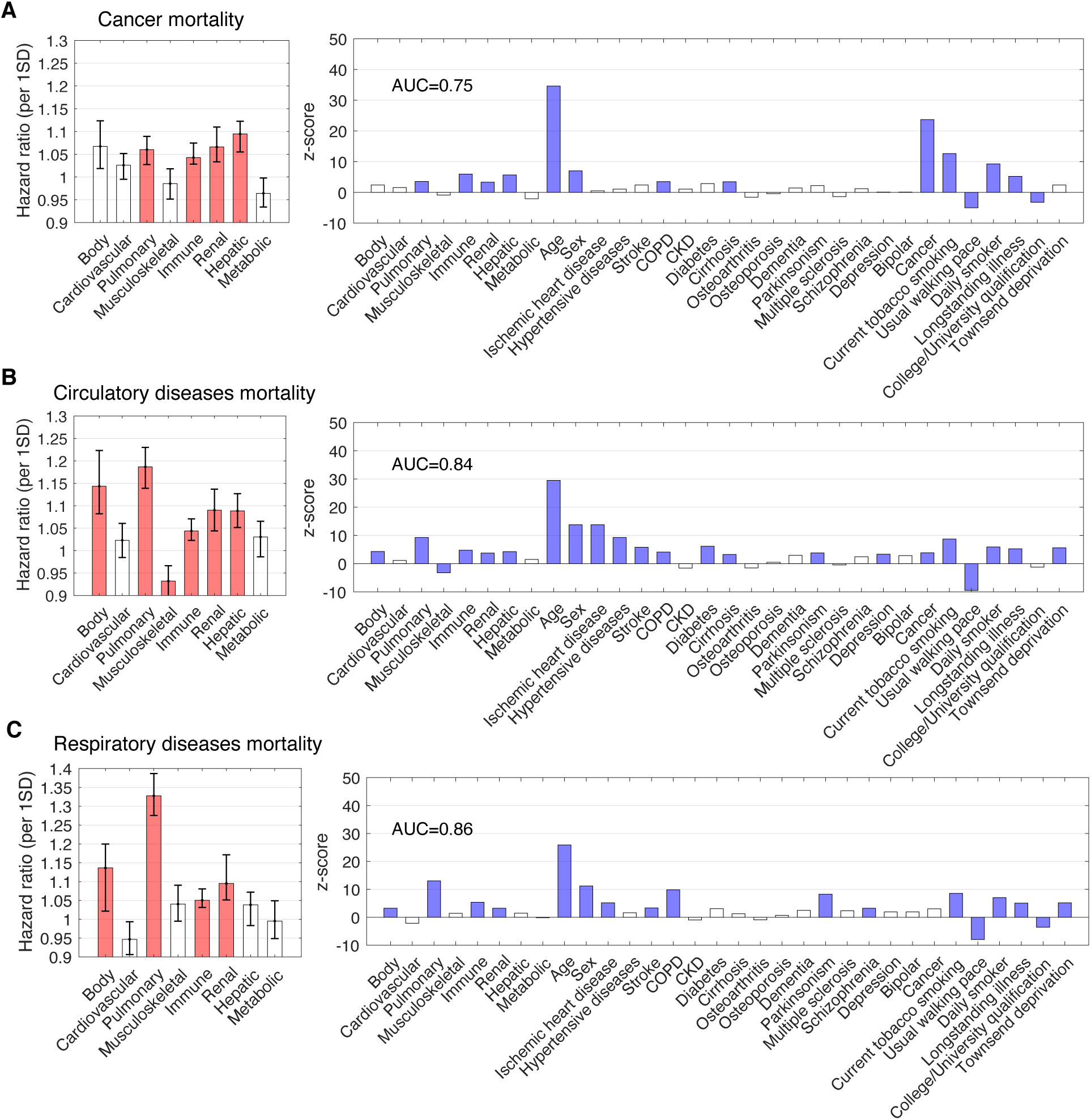
Body age and the risk of mortality. Mortality hazards for the three main causes of death, including cancer (**A**), circulatory diseases (**B**) and respiratory diseases (**C**). Bar plots show mortality hazard ratios per one standard deviation (SD) change in organ-specific age gap (left) and corresponding z-scores (right). Hazard ratios estimated using Cox proportional hazard regression, where survival durations were right censored for surviving individuals. Chronological age, sex, existing diagnoses, general health and key lifestyle factors included as confounds. Confidence intervals (95%) estimated with bootstrapping (n=100). Colored bars indicate body systems with significant hazard ratios (p<0.0019, Bonferroni corrected for 32 dependent variables). AUC: area under curve.

**Fig. S16.**
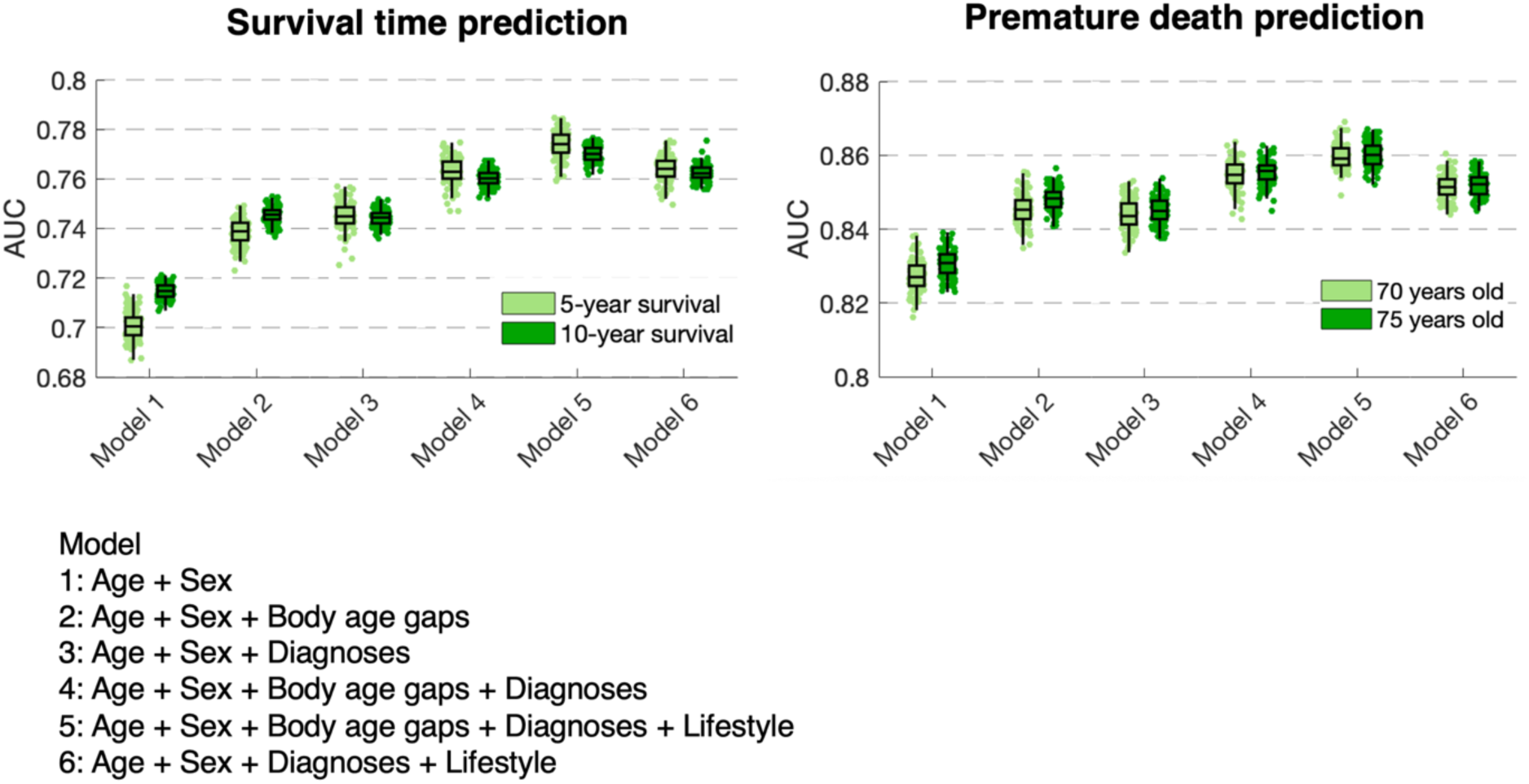
Survival time and premature death prediction. A logistic regression model was trained (10-fold cross-validation) to predict an individual’s 5- and 10-year survival (left) and premature death (defined as death before 70 or 75 years old, right). Boxplots show prediction accuracy, as quantified with area under curve (AUC). A hierarchy of six logistic models was established to determine the extent to which biological age improves prediction of survival time and premature death above and beyond established predictors (i.e., chronological age, sex, diagnoses, lifestyle factors). For prediction of both survival time and premature death, the model including body age gaps (Model 2) significantly outperforms the model including only chronological age and sex (Model 1, p<0.01). Similarly, the model including body age gaps (Model 4) significantly outperforms the model with only chronological age, sex and existing diagnoses (Model 3). Nevertheless, Model 5, which includes all predictors, achieves the most accurate predictions of survival time (5-year: AUC=0.774 ± 0.006; 10-year: AUC=0.770±0.003) and premature death (70 years old: AUC=0.86±0.003; 75 years old: AUC=0.86±0.003). Omitting body age gaps (Model 6) leads to significantly reduced accuracy (p<0.01) for predictions of survival time (5-year: AUC=0.76 ± 0.005; 10-year: AUC=0.76±0.003) and premature death (70 years old: AUC=0.85±0.003; 75 years old: AUC=0.85±0.003). Confidence intervals for AUC estimated with bootstrapping (100 samples).

**Fig. S17.**
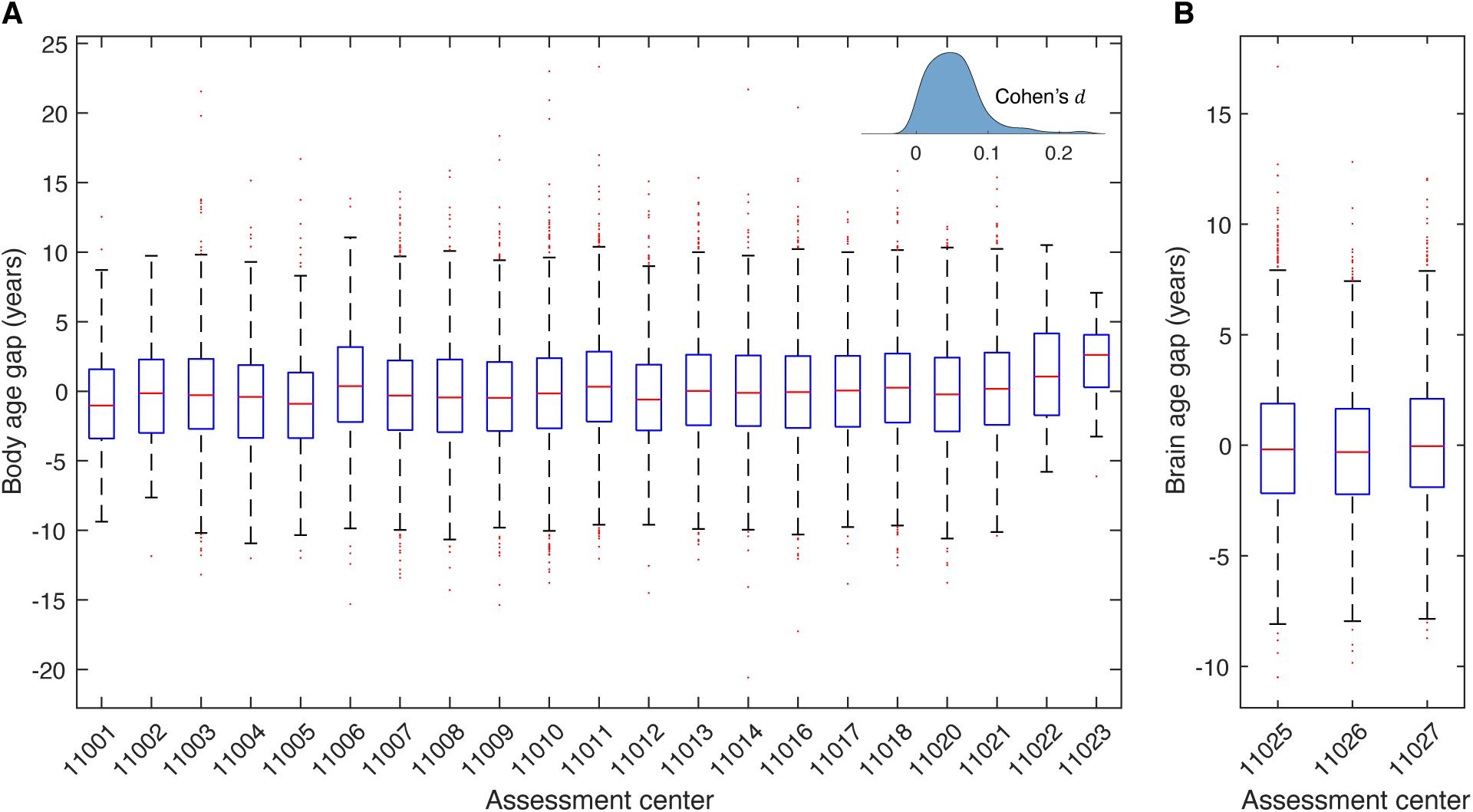
Site effects on estimated body and brain age. Body phenotypes were assessed at 21 different assessment centers, whereas brain images were acquired at 3 different centers. (**A**) Distribution of estimated body age gap across healthy aging individuals assessed in each of the 21 centers. The bottom and top edges of the boxes indicate the 25th and 75th percentiles of the distribution, respectively. The central red line indicates the median. The whiskers extend to the most extreme data points that are not considered outliers (1.5-times the interquartile range). Outliers are indicated with red dots. After regressing out sex and chronological age, one-way analysis of variance (ANOVA) shows that the differences in the mean of estimated body age gap across the 21 centers is significant (*F* statistic=8.98, p=1.35×10^-27^), but with a small effect size (Cohen’s 𝑓 =0.079). Inset shows the distribution of values of Cohen’s 𝑑 effect size (0.05±0.04) across 210 unique pairs of comparisons (two-sample t-test). (**B**) Distribution of estimated brain age gap across healthy individuals assessed in each of the 3 brain imaging centers. After regressing out sex and chronological age, one-way ANOVA shows that the differences in the mean of estimated brain age gap across the three centers is significant (*F* statistic=4.67, p=0.0094), but with a small effect size (Cohen’s 𝑓 =0.034). The values of Cohen’s 𝑑 effect size for the three pairs of comparisons (two-sample t-test) are 0.022, 0.024 and 0.05.

**Fig. S18.**
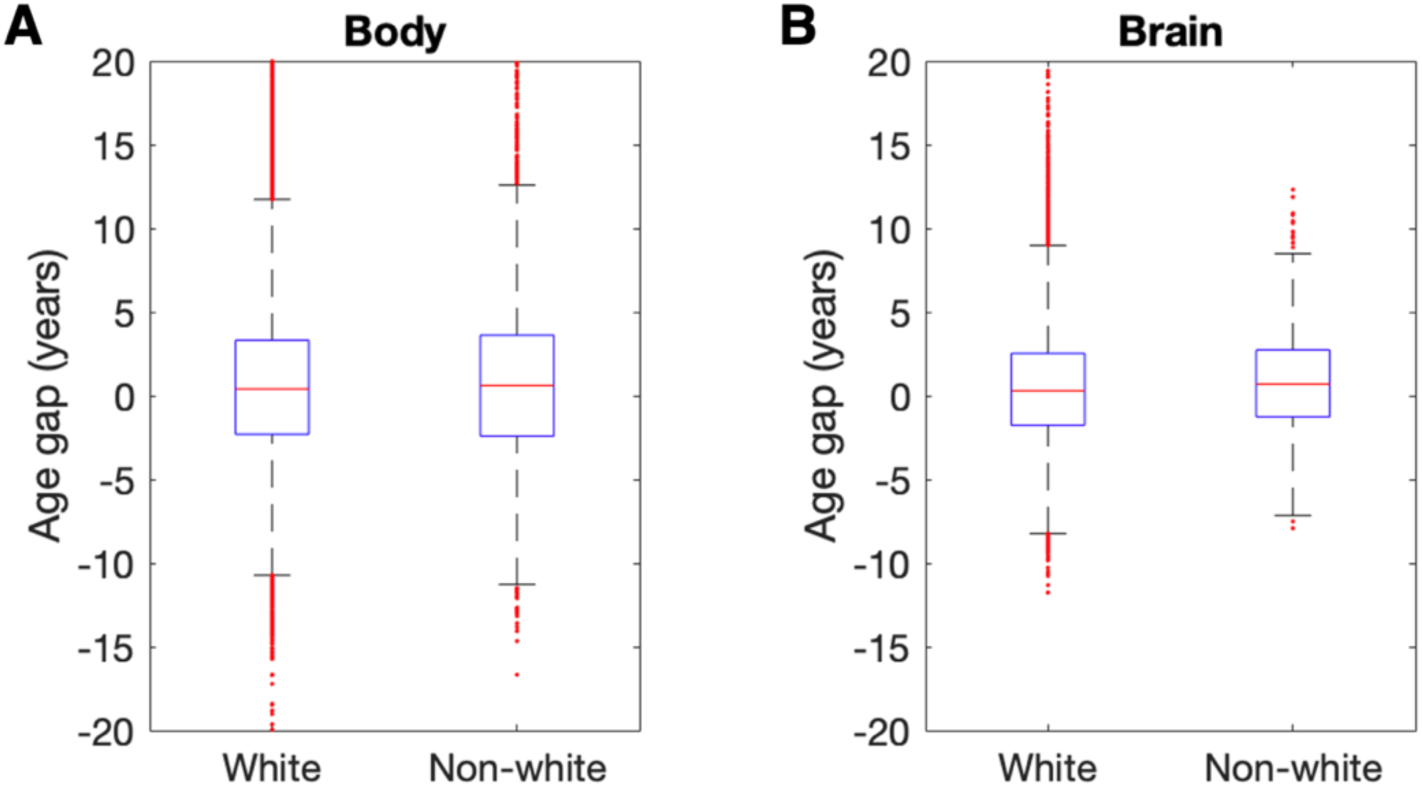
Ethnicity related variation in body and brain age. **(A)** Distribution of estimated body age gap across individuals from white (n=136,017, mean age gap=0.68±4.59 years) and non-white (n=7,406, mean age gap=0.89±5.20 years) ethnic backgrounds. Adjusting for chronological age and sex, the difference in the mean of estimated body age gap between white and non-white group is statistically significant (*t*=4.9, *p*=6.5×10^-7^), but with a small effect size (Cohen’s *d*=0.01). **(B)** Distribution of estimated brain age gap across individuals from white (n=35,704, mean age gap=0.48±3.33 years) and none-white (n=1,197, mean age gap=0.81±3.12 years) ethnic backgrounds. Adjusting for chronological age and sex, the difference in the mean of estimated brain age gap between white and non-white group is statistically significant (*t*=4.6, *p*=4.4×10^-6^), but with a small effect size (Cohen’s *d*=0.02). The bottom and top edges of the boxes indicate the 25th and 75th percentiles of the distribution, respectively. The central red line indicates the median. The whiskers extend to the most extreme data points that are not considered outliers (1.5-times the interquartile range). Outliers are indicated with red dots.

**Fig. S19.**
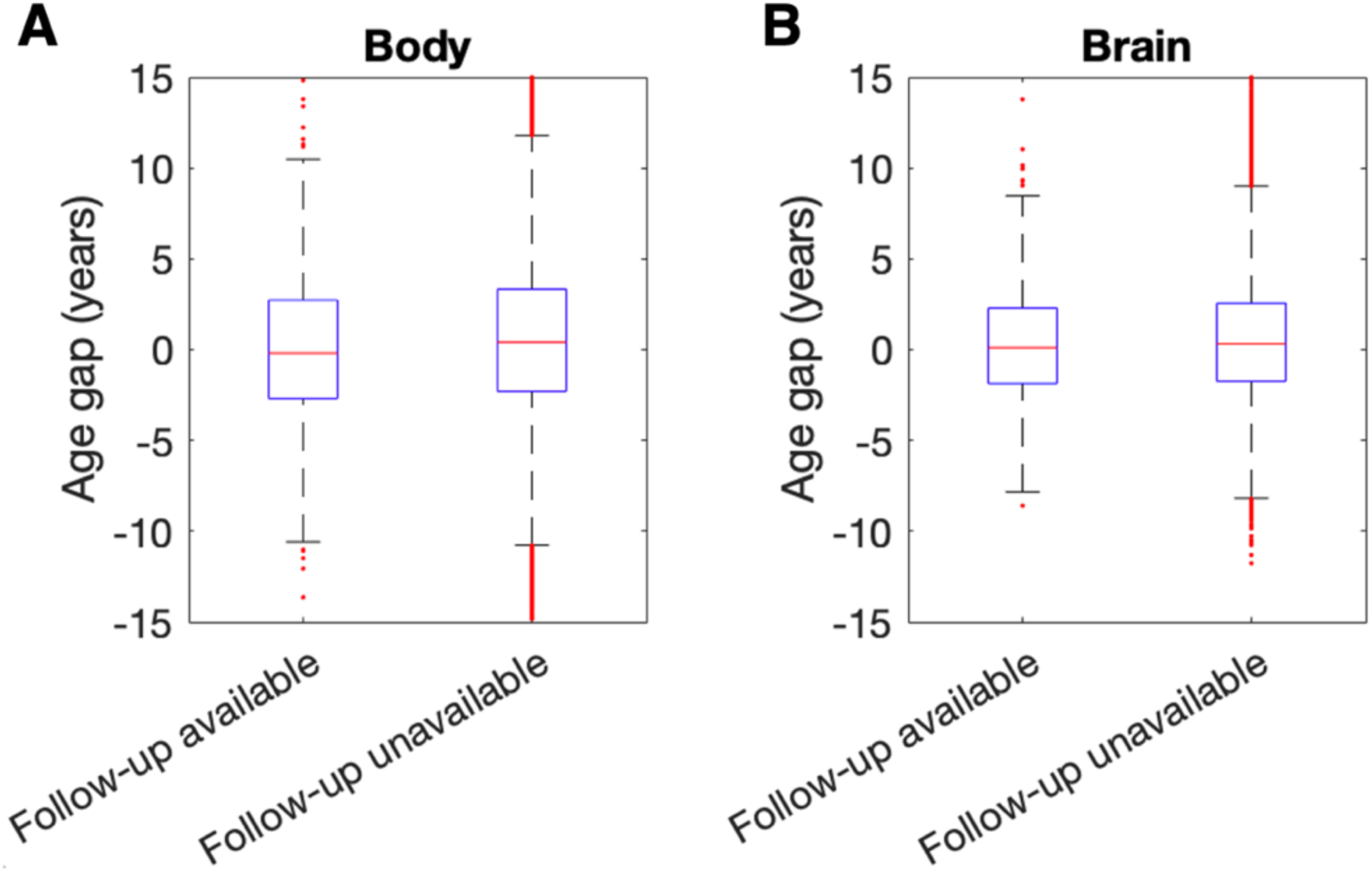
Subsampling related variation in body and brain age. **(A)** Distribution of estimated baseline body age gap across individuals with (n=1,220, mean age gap=0.005±4.25 years) and without (n=142,203, mean age gap=0.70±4.63 years) follow-up assessment. Adjusting for chronological age and sex, the between-group difference in the mean of estimated body age is significant (*t*=5.62, *p*=1.8×10^-8^), but with small effect size (Cohen’s *d*=0.015). **(B)** Distribution of estimated baseline brain age gap across individuals with (n=1,294, mean age gap=0.22±3.11 years) and without (n=35,607, mean age gap=0.50±3.33 years) follow-up assessment. Adjusting for chronological age and sex, the between-group difference in the mean of the estimated brain age gap is significant (*t*=2.53, *p*=0.01), but with small effect size (Cohen’s *d*=0.013). The bottom and top edges of the boxes indicate the 25th and 75th percentiles of the distribution, respectively. The central red line indicates the median. The whiskers extend to the most extreme data points that are not considered outliers (1.5-times the interquartile range). Outliers are indicated with red dots.

**Fig. S20.**
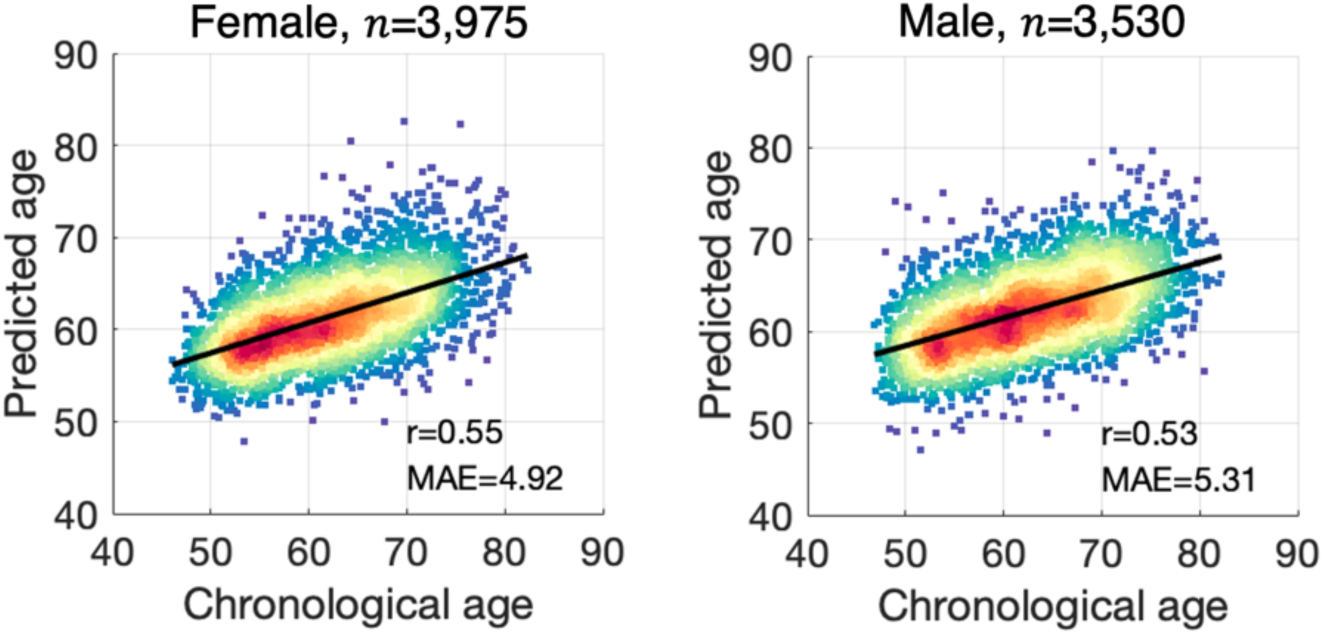
Age prediction using phenotypes derived from heart MRI and carotid ultrasound images. Scatter plots show associations between chronological age and predicted age for prediction model based on 28 imaging-derived phenotypes indexing cardiac function, blood pressure and carotid intima-medial thickness (table S14). For heart MRI, phenotypes measured during pulse wave analysis with plausible Vicorder results were included. Phenotypes derived from the inline VF (ventricle function) were not included due to the lack of quality control. Lines of best fit indicated with solid black lines. n: training sample size; r: Pearson correlation coefficients; MAE: mean absolute error.

## Captions for Supplementary Tables

**Table S1.**

Body age phenotypes.

**Table S2.**

Brain age phenotypes.

**Table S3.**

Cognitive phenotypes.

**Table S4.**

Model performance of each aging clock

**Table S5.**

Estimates from structural equation models for multiorgan aging networks.

**Table S6.**

Associations between genetic, environmental/ lifestyle factors and biological organ age.

**Table S7.**

Diagnostic codes related to the 16 disease categories.

**Table S8.**

Demographic details of individuals comprising the 16 clinical groups.

**Table S9.**

All-cause mortality risk estimates of body age gaps.

**Table S10.**

All-cause mortality risk estimates of brain age gaps.

**Table S11.**

All-cause mortality risk estimates of conventional body phenotypes.

**Table S12.**

All-cause mortality risk estimates of conventional brain phenotypes.

**Table S13.**

Cause-specific mortality risk estimates of body age gaps.

**Table S14.**

Imaging-derived phenotypes indexing cardiac and carotid function.

